# Recurrent Physical Contact Network Relevant for Epidemic Spreading

**DOI:** 10.64898/2026.04.08.26350390

**Authors:** Michael Lazarus Smah, Anna C. Seale, Kat S. Rock

## Abstract

Network-based epidemic models have been instrumental in understanding how contact structures shape infectious disease dynamics, yet widely used network modelling frameworks do not explicitly capture the combination of full (saturated) within-group interactions and constrained between-group links characteristic of many real-world settings. Here, we introduce the Multi-Clique (MC) network model to characterise recurrent physical contact networks, a framework in which individuals are organised into fully connected cliques representing stable contact groups (e.g., households, classrooms, or workplaces), with a limited number of external connections governing inter-group transmission. Using stochastic susceptible–infectious–recovered (SIR) simulations on degree-matched networks, we compare epidemic dynamics on MC networks with those on the Erdős–Rényi model, the configuration-model, and stochastic block networks. Despite having an identical mean degree, MC networks exhibit systematically distinct behaviour, including slower epidemic growth, reduced peak prevalence, increased fade-out probability, and delayed time to peak. These effects arise from rapid within but constrained between clique transmission, creating structural bottlenecks that standard models do not capture. By isolating the role of intergroup connectivity, the model offers a basis for evaluating targeted intervention strategies that reduce between-group mixing while preserving within-group interactions. Our results highlight the importance of explicitly representing real-life, clique-based network structure in epidemic models and suggest that classical degree-matched networks may systematically overestimate epidemic speed and intensity in structured populations.

## Introduction

Network models have become a foundational tool for understanding the spread of infectious pathogens, providing a principled way to encode heterogeneity in contact structure, transmission pathways, and population mixing patterns [13, 19], while relaxing the random-mixing assumption underlying classical equation-based epidemic models. By representing individuals as nodes and potentially infectious contacts as edges, network-based approaches have revealed how non-random mixing shapes epidemic thresholds, outbreak variability, and the effectiveness of different intervention strategies.

Network models in epidemiology consist of nodes representing individuals and edges representing *potentially infectious contact opportunities*, together with rules governing transmission along those edges. Epidemic processes such as susceptible–infectious–recovered (SIR) dynamics are then implemented on the network, with transmission occurring probabilistically when contacts are established between infected and susceptible nodes and recovery occurring at the node level. Within this framework, epidemic outcomes depend not only on pathogen-specific characteristics but also on structural properties of the network, including degree distributions, clustering, path lengths, community structure, and the presence of bottlenecks or highly connected individuals. These features jointly shape epidemic thresholds, growth rates, final outbreak sizes, and variability across different population groups. In static network models, edges represent persistent opportunities for repeated contact, whereas temporal network models resolve the timing and ordering of interactions; the appropriate description depends on whether epidemic dynamics are dominated by long-lived structural constraints or fine-scale temporal variation.

Classical random graph frameworks have played a central role in representing and analysing such network structures. Erdős–R’enyi (ER) random graphs [7, 8] serve as comparison models for homogeneous mixing and remain widely used as analytical benchmarks in epidemic theory [1, 13, 19]. The configuration model (CM) extends this framework by preserving empirical degree sequences, so that highly connected individuals and less connected individuals retain their observed contact frequencies even though the precise pattern of connections is randomised. This enables the study of degree heterogeneity, superspreading, and stochastic outbreak dynamics [20, 18]. Scale-free networks [4] further highlight the role of hubs and heavy-tailed degree distributions in lowering epidemic thresholds. The stochastic block model (SBM) encodes population structure through group-dependent mixing probabilities [9] and has been applied to model demographic, institutional, and spatially structured contact patterns in epidemic scenarios [3]. By tuning within- and between-group connection probabilities, SBMs capture heterogeneity in mixing rates across population subgroups.

Importantly, most network models now widely used in epidemiology were not originally developed to represent the potentially recurrent, physical contact processes characterising the household and work/school contact networks. Erdős–Rényi graphs were introduced as mathematical models of randomness, configuration models to generate graphs with prescribed degree sequences, scale-free networks to explain heavy-tailed connectivity patterns, and stochastic block models to identify community structure. These frameworks, focusing primarily on social network structures, have been adopted for epidemic modelling by interpreting edges as potential transmission pathways—an interpretation that has proven highly productive—but their principles remain largely different from mechanisms of repeated exposure, contact saturation, and stable group-based interaction.

Many human infectious diseases—particularly respiratory and close-contact infections—are spread primarily through *close contact*. In settings relevant to such transmission, contacts are predominantly recurrent rather than transient or randomly reassigned. Individuals tend to interact repeatedly with a small and relatively stable set of close associates, such as household members, classmates, coworkers, or patients within a ward. Within these settings, contacts are mostly dense and persistent, with everyone equally likely to meet with everyone else. Interactions outside the primary group may be unique to individuals and are comparatively sparse but still structured and may be recurrent. Within households, classrooms, or offices, individuals are likely to encounter most or all other group members over short time scales, whereas interactions between groups correspond to ongoing social ties rather than one-off encounters. Empirical contact studies consistently demonstrate strong group structure, repeated exposure, and limited mixing between groups [17, 6, 14, 16].

While classical network models are important, they do not explicitly encode the combination of near-complete exposure within small, stable groups and constrained inter-group contacts that characterise such environments. In the ER and configuration models, contacts are globally random once the degree constraints are fixed; in SBMs, within-group density is controlled probabilistically rather than representing fully realised, repeatedly accessible contact opportunities. As a result, these models do not directly capture the saturation of transmission pathways within groups or the limited inter-group connections governing between-group spread. Motivated by this structural gap, this study introduces the *Multi-Clique (MC) network model*, explicitly designed to represent full mixing opportunities within groups, reflecting physical contact networks. In the MC framework, individuals are partitioned into small, fully connected cliques representing stable contact groups. External connections to other cliques of any individual are independent of their remaining clique members. This construction directly encodes saturation within groups and constrained transmission pathways between groups, providing an interpretable abstraction of recurrent contact structure that characterises human daily interactions.

## Multi-Clique Network Model

### Definition

#### Definition 1

*Let N* ∈ N *denote the total number of nodes. Let S be a random variable representing clique sizes, with mean* 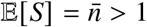, *and let* 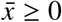 *denote the mean external degree*.

*An undirected graph G* = (*V, E*), *where V is the set of nodes and E is the set of edges, with* |*V* | = N, *is called a* multi-clique (MC) network *if it is constructed according to the following conditions:*

1. *The node set V is partitioned into K disjoint subsets (cliques), where K denotes the total number of cliques:*

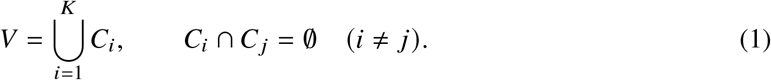

*The size of clique C*_*i*_ *is* |*C*_*i*_ | = *S*_*i*_, *where S*_*i*_ *is a realisation of the random variable S, and*

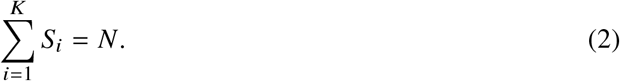

*2*.*Each clique C*_*i*_ *induces a complete subgraph. Hence, for all distinct nodes u, V* ∈ *C*_*i*_, *an edge exists between them, i*.*e*.,

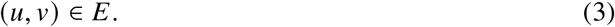

*3. Each node u* ∈ *V is assigned an initial external-degree target x*_*u*_ ∈ Z_≥0_, *drawn from a distribution with mean* 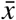.

*4. External half-edges are paired uniformly at random, subject to the constraint that external edges connect nodes belonging to different cliques. That is, if an external edge joins nodes u and v, then*

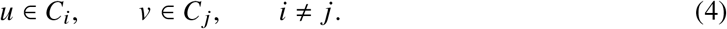

*To ensure connectivity between cliques, a repair step is subsequently applied. Specifically, if a clique C*_*i*_ *has no external edge following the pairing process, a single inter-clique edge is added between a node in C*_*i*_ *and a node belonging to another clique. Consequently, every clique is connected to at least one other clique, although the realised external degree sequence may differ slightly from the initially assigned external degree sequence*.

*Consequently, for every clique C*_*i*_, *there exists at least one node u* ∈ *C*_*i*_ *and one node v ∉ C*_*i*_ *such that* (*u, v*) ∈ *E*.

*The degree of a node u* ∈ *C*_*i*_ *is therefore*

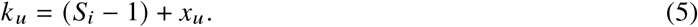

### Construction of MC Network

#### Stage 1

Generate clique sizes 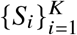 as realisations of a random variable *S* ~ *P*(*S*), subject to the constraint

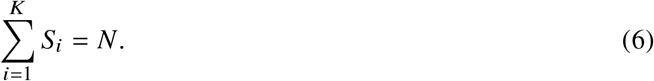

To achieve heterogeneous clique sizes, *P*(*S*) may be chosen from a suitable family of distributions, such as Poisson, geometric, negative binomial, or power-law distributions.

For Poisson-distributed clique sizes, we use a Poisson distribution truncated below at one. Specifically, clique sizes are generated as

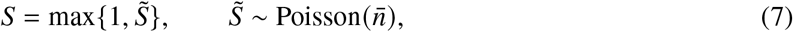

which permits singleton cliques while excluding empty cliques.

#### Stage 2

- Fully connect nodes within each clique, so that each node *u* ∈ *C*_*i*_ has intra-clique degree

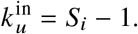
- Assign an external degree *x*_*u*_ to each node *u* ∈ *V*.
- Generate inter-clique edges restricted to pairs of nodes belonging to distinct cliques.

This construction enables independent control of clustering (via *P*(*S*)), connectivity 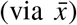, and heterogeneity (via both *P*(*S*) and the external-degree distribution).

### Mean Degree

The mean degree depends on whether clique sizes are homogeneous or heterogeneous.

#### Homogeneous clique sizes

If *S*_*i*_ *≡* 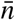 for all _*i*_, then each node has intra-clique degree 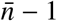. Hence, the mean degree is

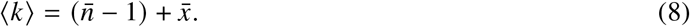

#### Heterogeneous clique sizes

Let *S* denote the clique-size random variable with distribution *P*(*S*). We distinguish between two sampling perspectives:

- *Clique-level sampling:* cliques are sampled with probability *P*(*S*).
- *Node-level sampling:* nodes are sampled uniformly at random.

Since a clique of size *s* contains *s* nodes, larger cliques are proportionally more likely to be sampled when selecting a node uniformly at random. Consequently, the clique size observed by a randomly chosen node follows the *size-biased distribution*

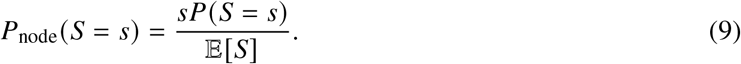

The expected clique size from the perspective of a randomly chosen node is

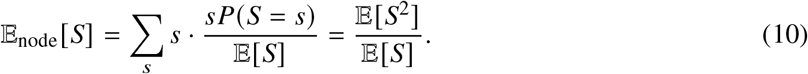

Each node in a clique of size *S* has intra-clique degree

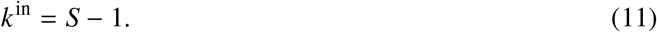

Therefore,

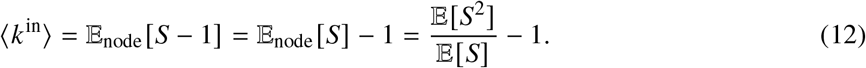

Adding the contribution from external edges, whose mean degree is 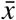, yields

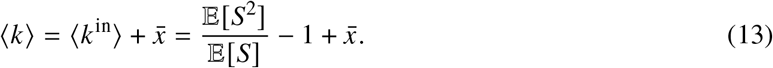

Using the identity

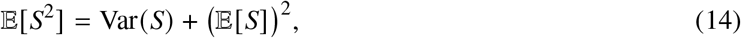

we obtain

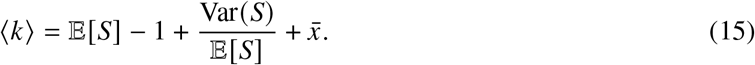

This implies that heterogeneity in clique sizes (Var(*S*) > 0) strictly increases the mean degree relative to the homogeneous case, reflecting the effect of size-biased sampling.

For illustration, suppose clique sizes follow a Poisson distribution with mean 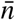, i.e.,

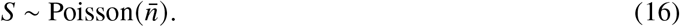

Then

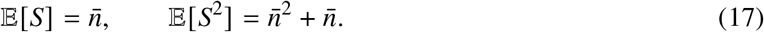

Substituting into (13) gives

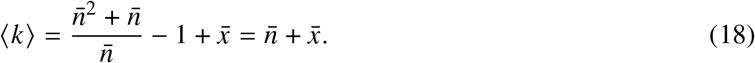

Thus, for Poisson clique sizes,

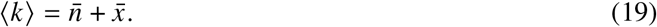

More supporting information for this work are available in the accompanying Supplementary Information (**SI). Algorithm 1** in the **SI** is a pseudocode, which provides a procedure for generating realisations of the MC network model. Figure 1 shows representative network realisations generated using the corresponding R implementation.

**Figure 1:**
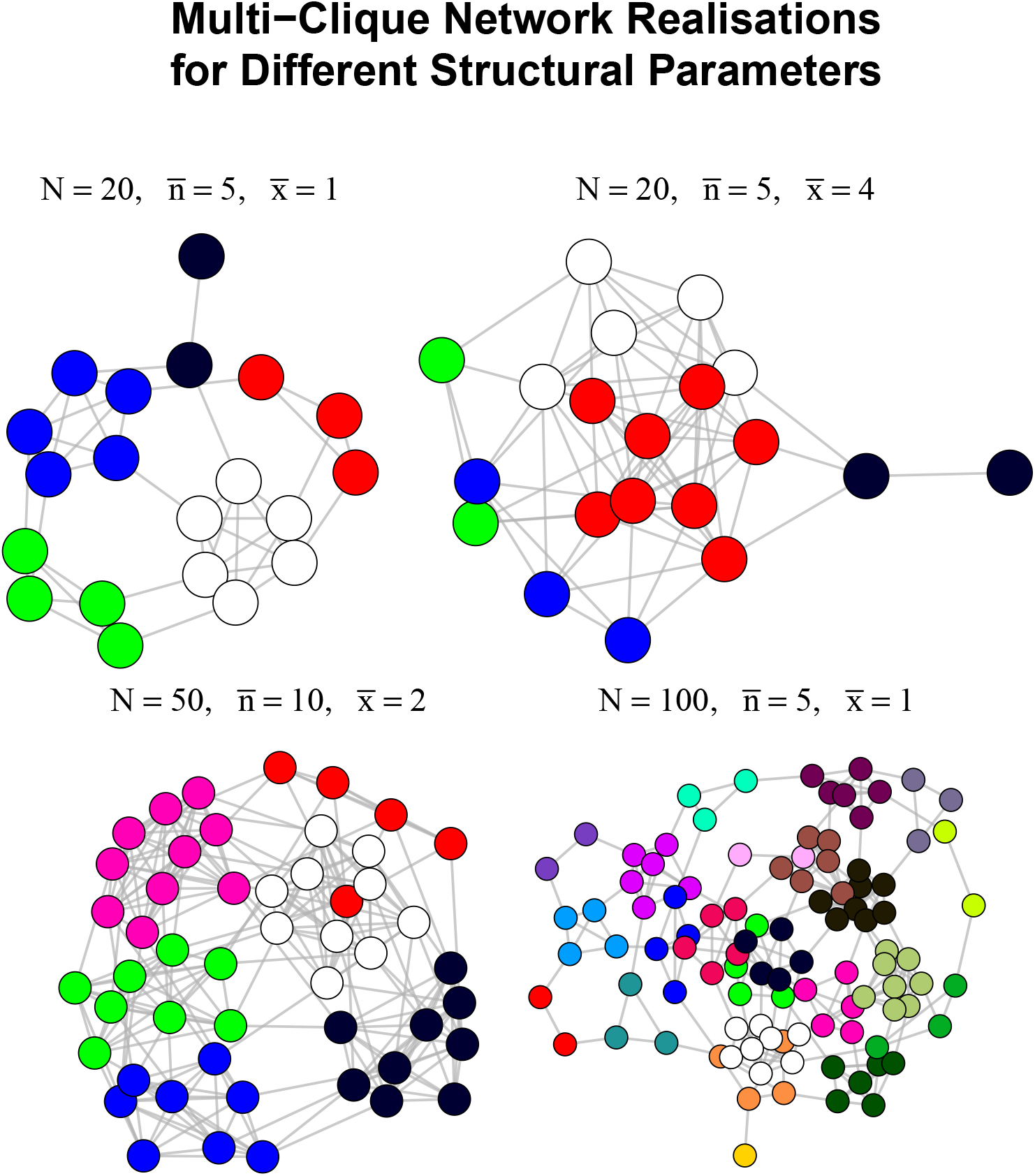
Illustrative realisations of the Multi-Clique (MC) network model generated using an R implementation of **Algorithm 1** (see **SI**) for different values of the population size *N*, mean clique size 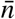, and mean number of external connections per individual 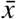. Each network is composed of fully connected cliques (shown in distinct colours), together with random inter-clique edges connecting individuals belonging to different cliques. Increasing N produces larger network structures, increasing 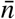 yields larger and fewer cliques, while increasing 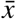 increases the density of inter-clique connectivity and strengthens mixing between cliques. The network with N = 100, 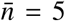, and 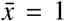 exhibits a large population organised into many small, tightly connected cliques with relatively few external links, whereas the network with *N* = 20, 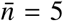, and 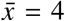 displays greater connectivity between cliques.

A practical advantage of the MC formulation is that its parameters admit direct empirical interpretation. Clique sizes correspond to stable contact groups such as households, classrooms, or work teams, while the mean external degree 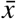 represents the number of potentially recurrent contacts outside the primary group. Both quantities can be estimated from administrative data and standard contact surveys (see Table 1) without requiring full network reconstruction.

**Table 1:** Interpretation and empirical grounding of Multi-Clique (MC) model parameters.

| MC parameter | Interpretable as | Possible data source |
| --- | --- | --- |
| $\bar{n}$ | Mean household / class / work-team size | Census data, school registers |
| $\{n_k\}$ | Distribution of group sizes | Administrative records |
| $\bar{x}$ | Mean number of external recurrent contacts | Contact surveys, diaries |
| Clique identity | Stable social affiliation | Household, workplace, ward data |

### Epidemic dynamics

In this study, epidemic spread is modelled using a discrete-time susceptible–infectious–recovered (SIR) process similar to the ones in the literature (see [1] for example). At each time step, infectious nodes transmit infection independently to each susceptible neighbour with probability *β*, and recover with probability *γ*. Simulations are initiated with a single randomly chosen infectious individual and run for a fixed number of time steps.

To assess the epidemiological consequences of the multi-clique structure, epidemic dynamics on MC networks are compared with the ER, CM, and SBM networks matched in population size and mean degree (average number of edges or connections per node). These models were selected for their distinct and well-understood structural mechanisms relevant to epidemic spread. ER networks serve as a baseline for homogeneous random mixing. CM networks preserve the full degree sequence of the MC networks, while they do not consider group saturation. SBMs encode a community-style network; however, they do not enforce the deterministic intra-group connectivity of the MC network. Importantly, these models are widely used, analytically interpretable, and generate static contact networks in which edges represent opportunities for transmission, matching the modelling assumptions of the MC framework.

For each choice of the MC parameters 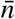, 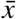, we constructed the MC network. The mean degree ⟨*k*⟩ of the generated MC network is computed. This process is to address the possibility that ⟨*k*⟩ may not always be exactly the same as 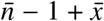 due to heterogeneity in clique sizes. All comparison networks are then generated with the same number of nodes N and matched mean degree ⟨*k*⟩. This degree-matching procedure ensures that differences in epidemic behaviour reflect structural features—such as clustering and clique structure—rather than differences in population size and number of contacts.

All epidemic simulations are performed on static networks, which are held fixed for the duration of each outbreak and regenerated only between independent realisations. As a result, all networks considered (MC, ER, CM, and SBM) represent *potentially-recurrent* contact structures in the sense that each node can only be infected by the pre-assigned neighbours, as edges encode persistent opportunities for repeated contact over time.

For each network type and parameter setting, epidemic trajectories are averaged over 10, 000 independent realisations of both the network and the stochastic SIR process. The mean number of infectious individuals over time is recorded. To characterise the structural features underlying epidemic dynamics, standard network statistics, including the mean degree ⟨*k*⟩, and the average local clustering coefficient, defined as the mean of node-wise clustering coefficients is computed.

The ER, CM, and SBM networks are constructed using the standard igraph package in R [5] with parameters chosen to match N and ⟨*k*⟩, while MC networks are generated using **Algorithm 1** (**SI**), yielding an adjacency matrices used for simulating outbreaks. All networks are considered to be undirected, meaning that if an infected node _*i*_ can infect a susceptible *j*, then an infected *j* can infect a susceptible _*i*_. The computed quantities, the mean degree ⟨*k*⟩, and the average local clustering coefficient are averaged over independent realisations to account for stochastic variability in both network generation and epidemic dynamics.

All simulations are performed on networks of fixed size N = 1, 000. For the MC model, the mean clique size is varied, such that 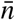 ∈ {1, 2, 4, 5, 10, 20}, while the mean external degree 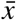 is varied over the range 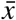. Epidemic dynamics follow a discrete-time SIR process with transmission probability *β*= 0.25, recovery probability γ= 0.10, and a maximum simulation time of *T*_max_ = 60 time steps. Each simulation is initialised with a single randomly chosen infectious node. For each pair of values 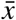,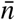 the empirical mean degree ⟨*k*⟩ of the MC network is first computed and averaging over 10, 000 MC realisations. Comparison networks are then generated with the same number of nodes N and matched mean degree ⟨*k*⟩. The ER networks are generated using the *G*(*N, p*) model with *p* = ⟨*k*⟩/(*N* − 1). The CM networks are generated by drawing random graphs with degree sequence with mean ⟨*k*⟩. Stochastic block models (SBMs) are constructed with block sizes of approximately 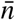, a fixed within-block connection probability *p*_in_ = 0.5, and a between-block probability *p*_out_ chosen so that the resulting mean degree matches ⟨*k*⟩. For each network model and parameter setting, epidemic trajectories and network statistics are averaged over 10, 000 independent realisations of both the network and the stochastic SIR process, ensuring robustness that account for intrinsic stochastic variability.

## Results

Supplementary Figures (in the **SI**) 1–6 show that the realised mean degrees are closely matched across network models. For most 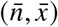 combinations, ER, MC, and SBM exhibit overlapping distributions of realised mean degree, while CM produces slightly higher realised mean degrees when 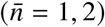. These differences arise because degree matching is imposed in expectation, whereas the realised mean degree varies slightly across stochastic network realisations. For 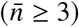, the realised mean degrees of all four models differ by less than one on average. At 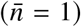, the MC model reduces to a degree-controlled random graph and the realised mean degrees overlap almost completely across models. As 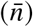 increases, the expected mean degree rises through the deterministic intra-clique contribution 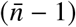 together with the external connectivity parameter 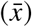. We therefore achieve close matching of average degree across models, while preserving differences in higher-order structural properties such as clustering and community organisation.

Supplementary Figures 7–12 showing local clustering reveal the core structural distinction. MC networks exhibit substantially higher average local clustering coefficients than the comparator models for all 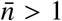, with the gap widening dramatically as 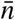 increases. For 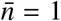, clustering is low (below 0.1) across all four models, although MC shows visibly higher clustering coefficient. For 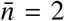 clustering in MC is already markedly elevated; by 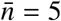 the clustering coefficient reaches 0.4–0.6 depending on 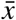, and for 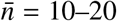 it approaches 1. In contrast, ER, CM, and SBM clustering remains near zero (typically < 0.05) across the entire range of 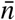 and 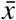 Within MC, clustering decreases with increasing 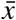 once 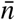 is fixed.

Figures 2–4, and Supplementary Figures 19–24 show that epidemic trajectories differ systematically once clique structure is present. At 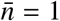 all four models produce nearly indistinguishable mean infection curves for every 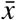: rapid rise, similar peak height and timing, and comparable decay. As 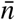 increases, the MC trajectories diverge. For 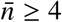 (especially at low-to-moderate 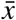), MC outbreaks show delayed take off, lower peak prevalence, and slower overall growth relative to ER, CM, and SBM. The delay is most pronounced at small 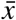, where infection often saturates the initial clique before inter-clique transmission occurs; higher 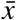 allows earlier escape, but MC still lags the benchmarks. ER and CM curves remain fast and similar to each other; SBM is intermediate but faster than MC because probabilistic within-block mixing avoids full local saturation. At high 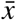 (5–6) and large 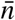, the qualitative shapes converge somewhat, but MC peaks remain delayed. When plotted per network type (Supplementary Figures 19–24), MC curves at low 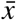 are visibly flattened and right-shifted compared with the overlapping families of curves for the other three models.

**Figure 2:**
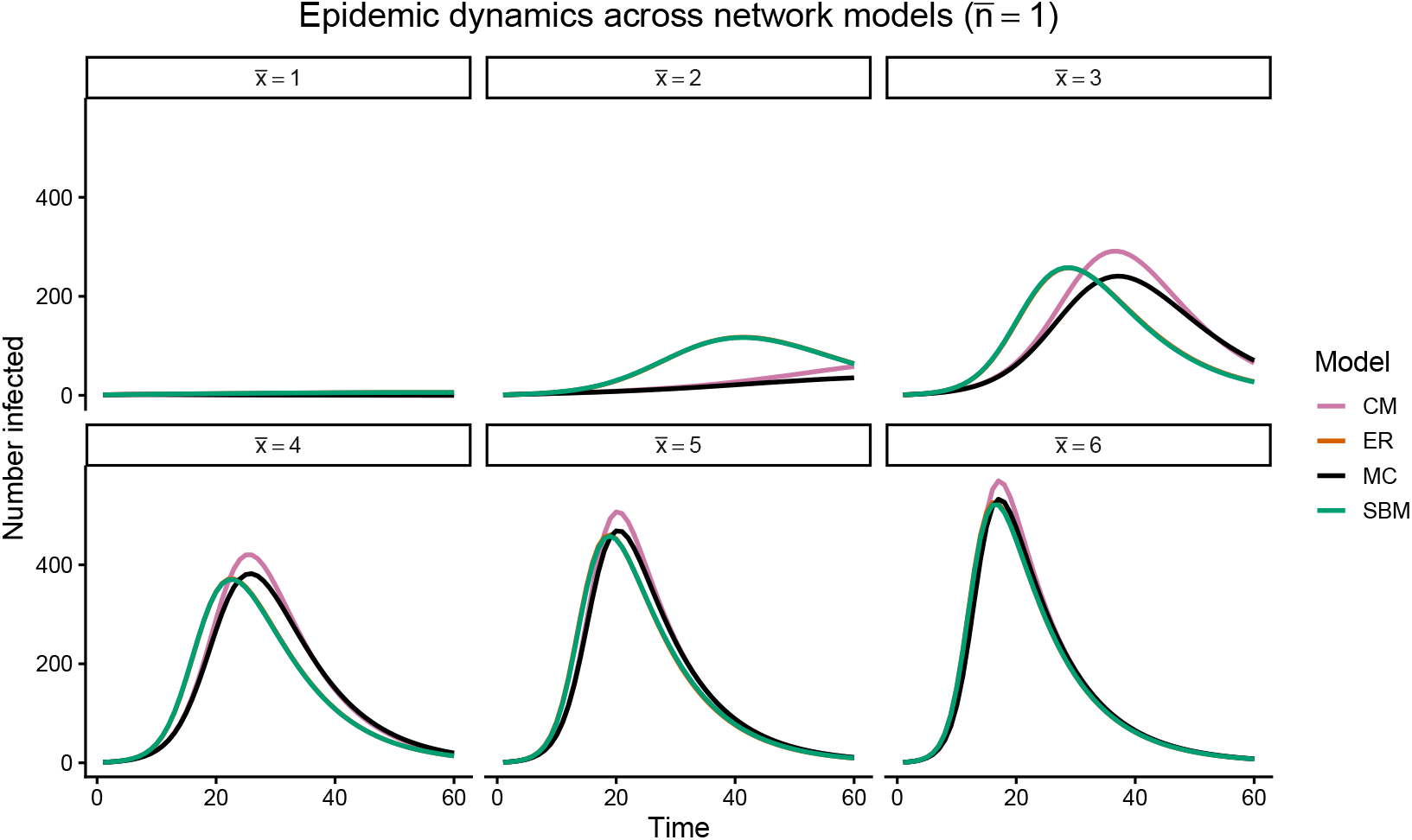
Network structure impact on epidemic trajectories for an average clique size of 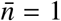. Each panel fixes the mean number of external contacts to 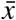. Curves show the average number of infections over time for an SIR model on the four different networks (CM, ER, MC, SBM) over stochastic simulations.

**Figure 3:**
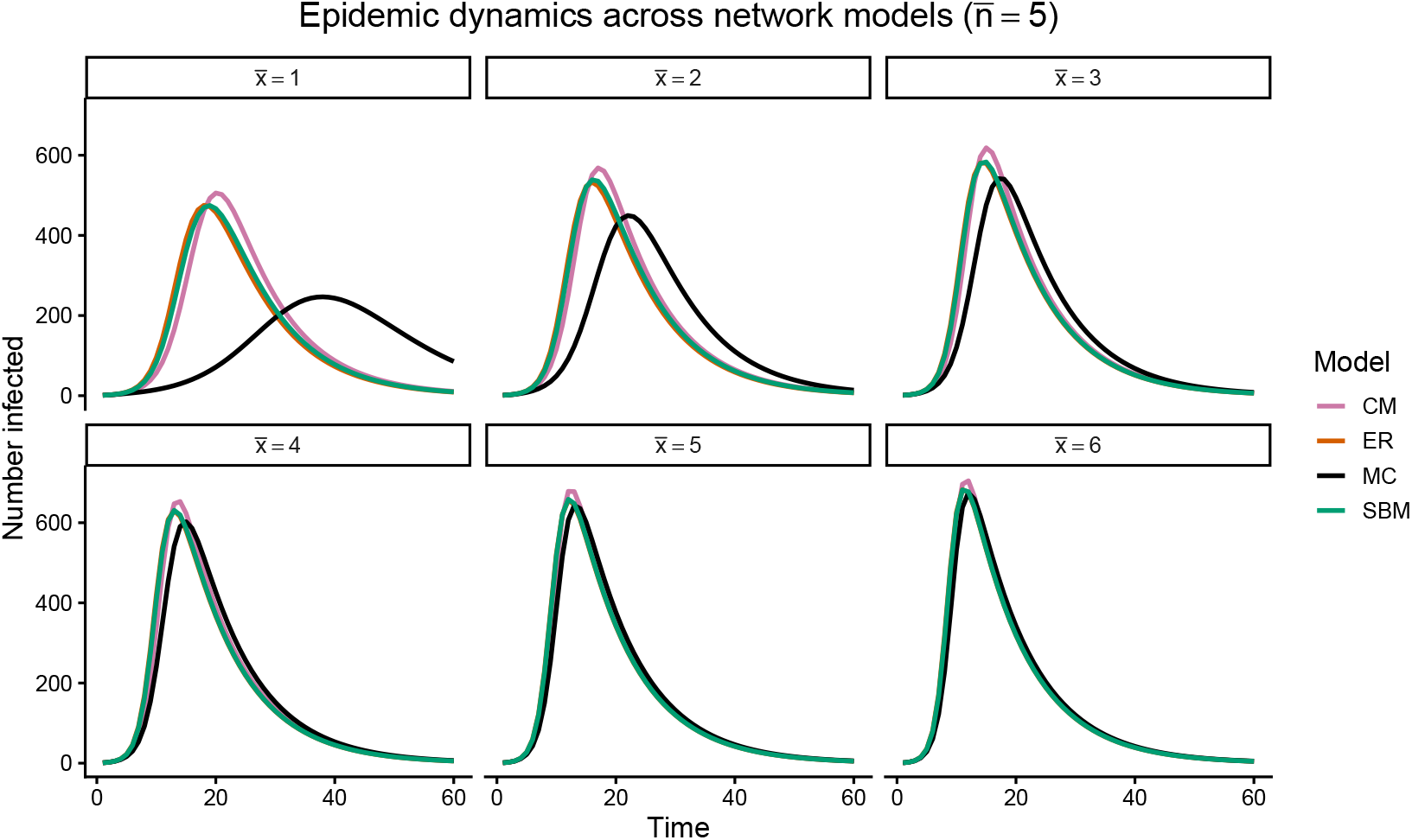
Network structure shapes epidemic trajectories. Mean SIR infection dynamics for 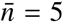 across network models. Each panel fixes external connectivity 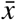. Curves show averages over stochastic simulations.

**Figure 4:**
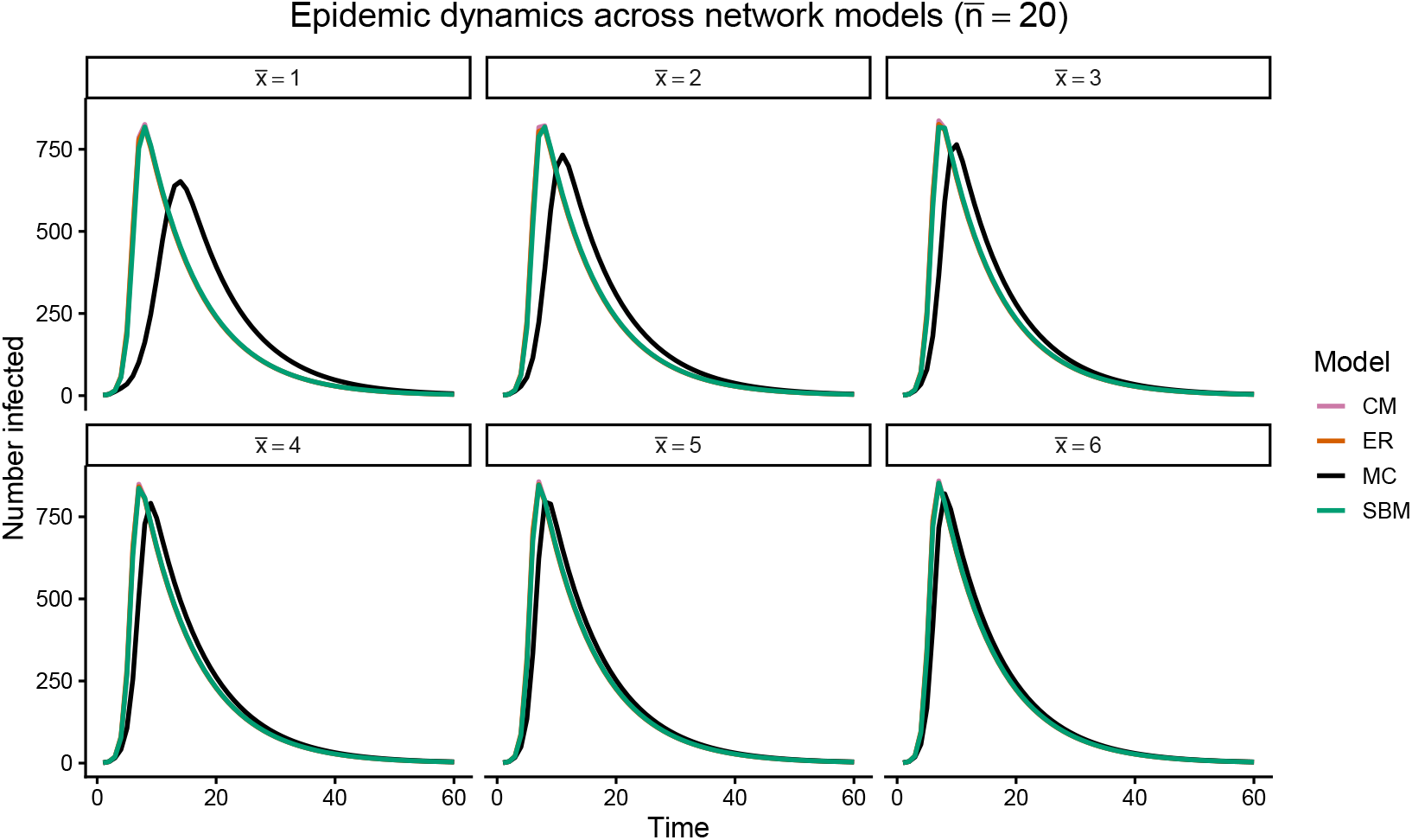
Network structure shapes epidemic trajectories. Mean SIR infection dynamics for 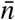 = 20 across network models. Each panel fixes external connectivity 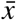. Curves show averages over stochastic simulations.

Figure 5 and Supplementary Figures 25–30 showing summary epidemic outcomes highlight the structural effects of the MC network compared to the other network models. Final epidemic size increases with 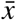 in all models and approaches similar values at high 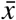, indicating agreement on ultimate reach once transmission percolates globally. However, MC consistently shows modestly smaller final sizes at intermediate 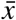, especially for larger 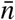. Peak prevalence follows the same pattern: comparable at 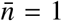 and high 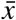, but MC exhibits lower peaks (sometimes substantially) for 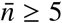 and 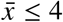. Fade-out probability is markedly higher on MC networks at low 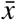, reflecting frequent extinction inside the seed clique. The time to peak is longer on MC across almost all regimes, with the largest differences at small 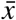 and moderate-to-large 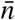; ER, CM, and SBM peak earlier. All models agree on the qualitative monotonicity (larger 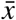 leads to bigger and faster outbreaks,lower fade-out, and earlier peaks), but disagree quantitatively once 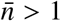, with MC systematically slower, lower-peaking, and more prone to stochastic extinction.

**Figure 5:**
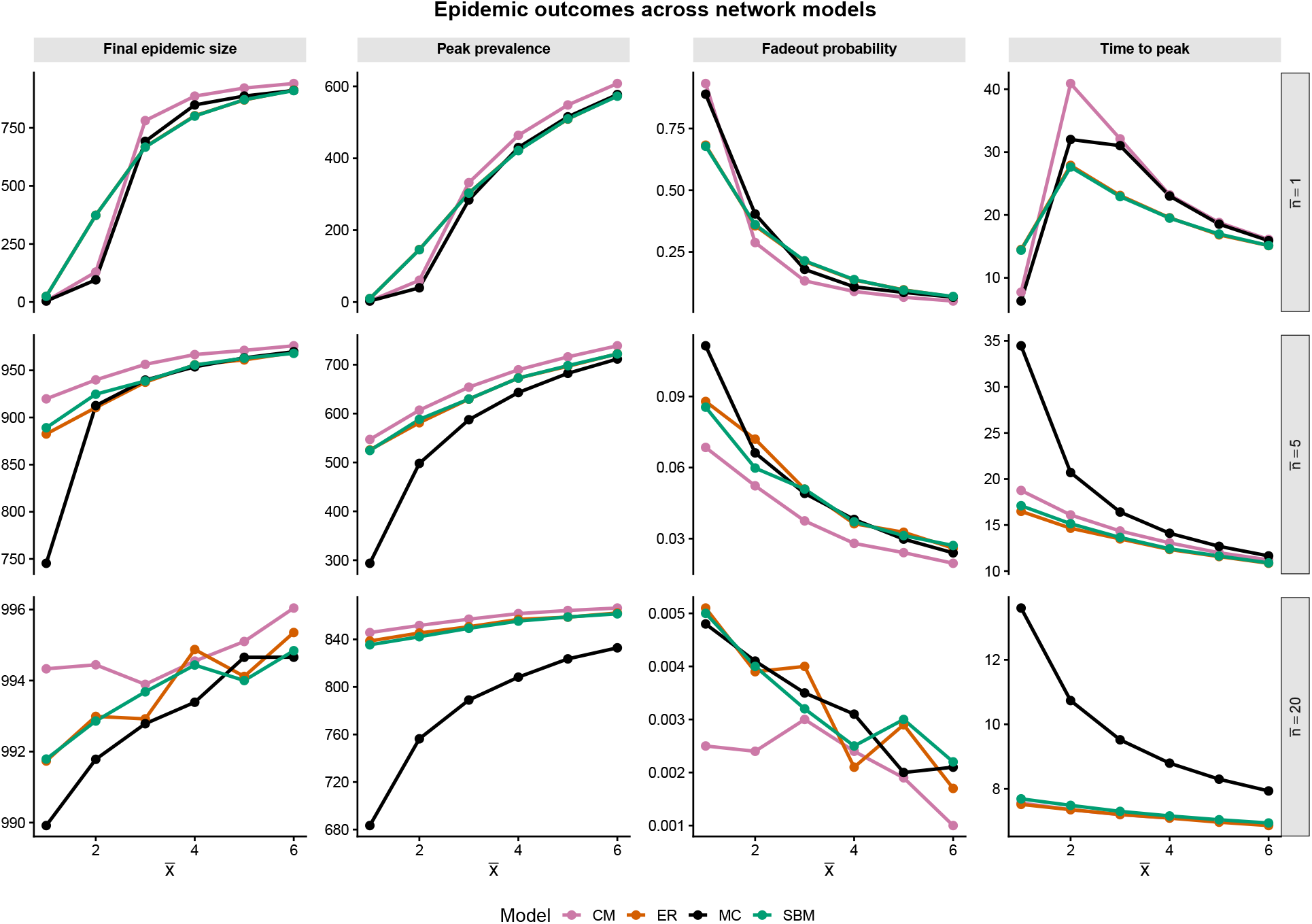
Comparison of epidemic outcomes across Erdős–Rényi (ER), stochastic block model (SBM), configuration model (CM), and multi-clique (MC) networks under SIR dynamics. Columns correspond to four epidemiological outcome measures measured across 10,000 realisations: final epidemic size, peak prevalence, fadeout probability, and time to epidemic peak. Rows correspond to different mean clique sizes, 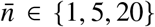 (indicated on the right-hand side), while the horizontal axis represents the mean external connectivity, 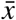 = 1 − 6.

Figures 6–8 show suppressed and delayed epidemics in MC networks relative to the comparator models, and the differences become more pronounced with increasing the mean clique size 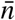. For fixed 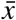, final epidemic size declines mildly with 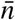on MC while remaining high and stable on ER/CM/SBM; the relative gap widens at lower 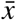. Peak prevalence decreases more noticeably with 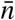 on MC than on the benchmarks, which maintain higher peaks. Time to peak shortens overall with 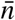(larger cliques accelerate local saturation), but MC still peaks later than the others for each fixed 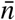 and 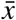. These trends reinforce that deterministic full connectivity within larger groups creates strong local bottlenecks that the probabilistic mixing of SBM (and the lack of explicit groups in ER/CM) do not replicate.

**Figure 6:**
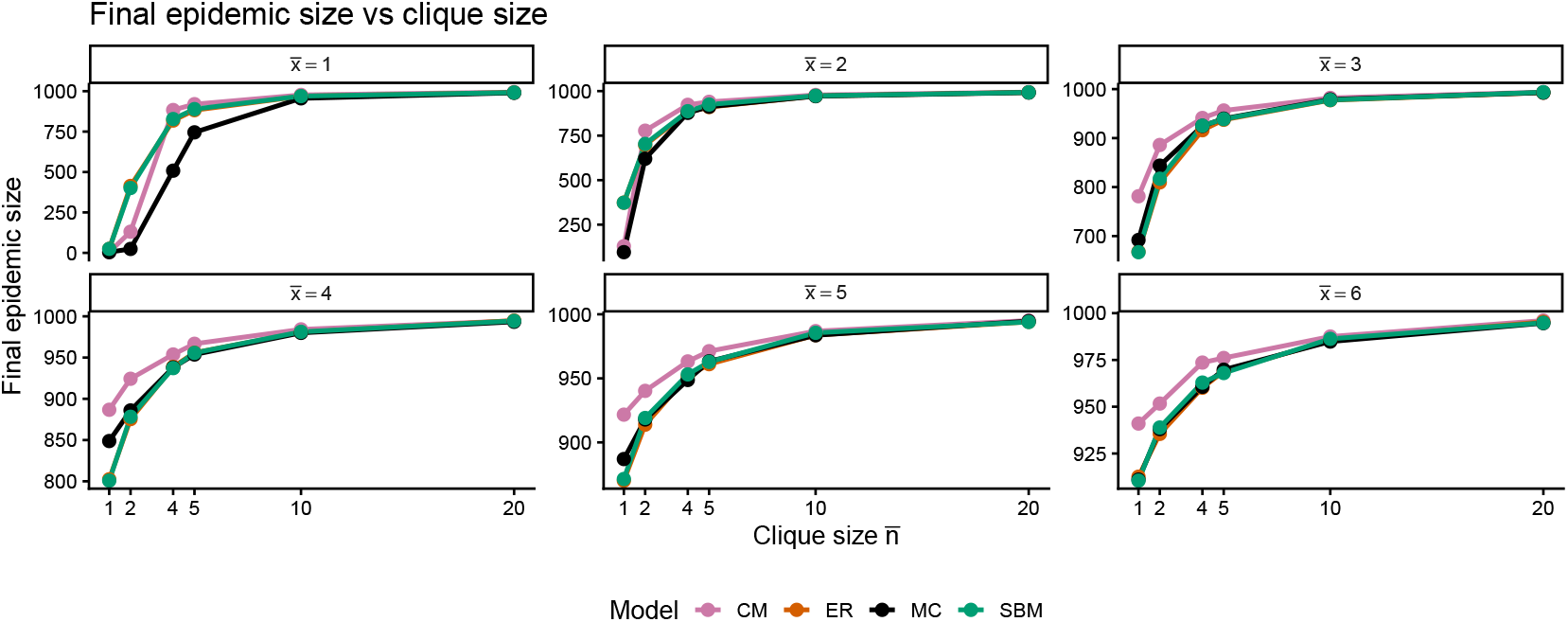
Clique size suppresses epidemic extent. Final epidemic size as a function of average clique size 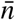. Each panel fixes external connectivity 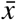. Increasing 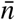 reduces outbreak size in multi-clique networks relative to degree-matched alternatives.

**Figure 7:**
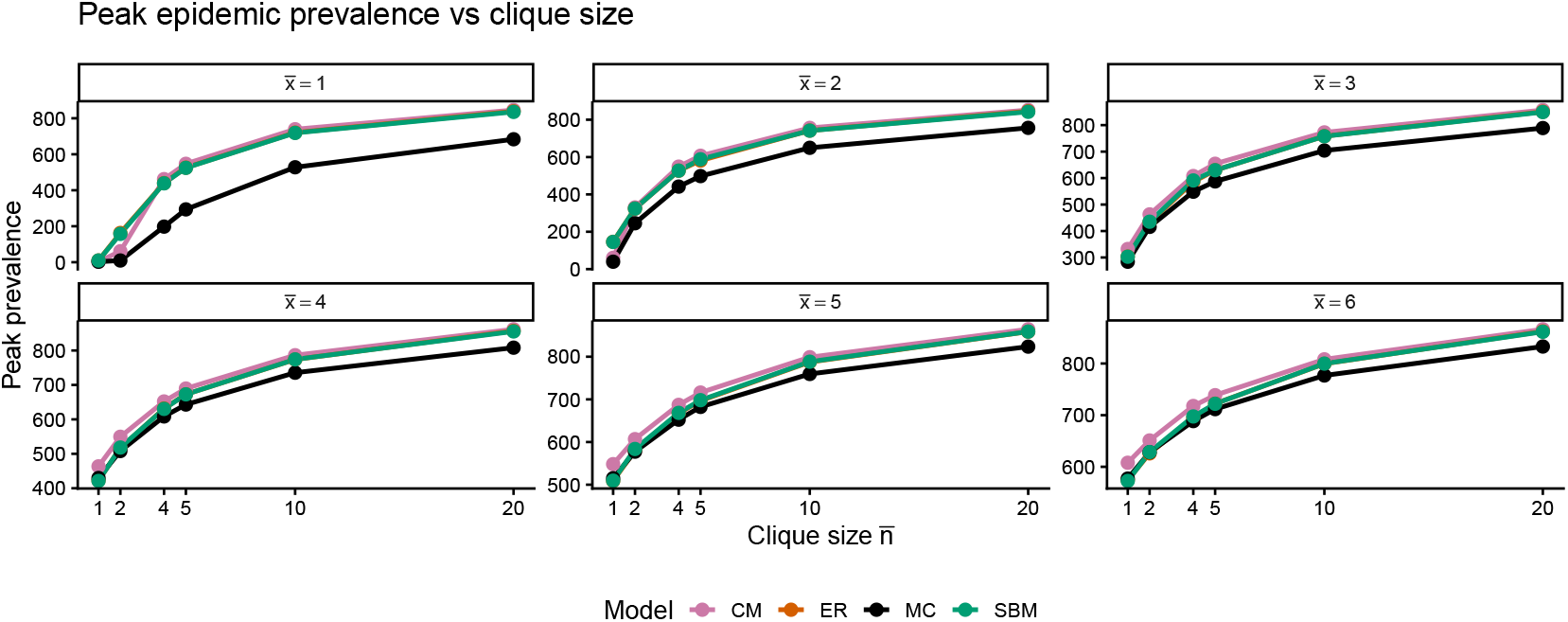
Peak prevalence decreases with clique size. Peak epidemic prevalence as a function of average clique size 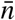 for fixed external connectivity 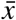. Multi-clique networks consistently exhibit lower peaks than comparing models.

**Figure 8:**
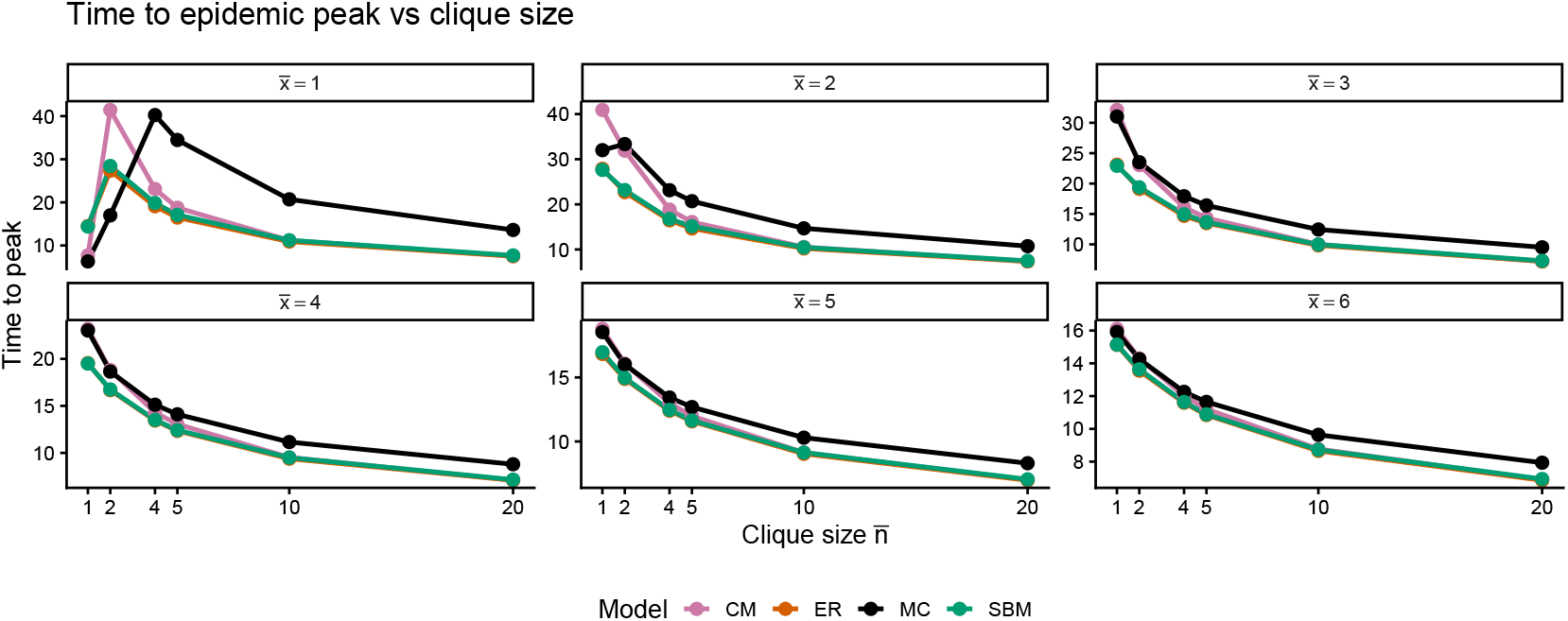
Clique structure delays epidemic spread. Time to epidemic peak as a function of average clique size 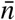. Each panel fixes external connectivity 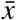. Epidemics on multi-clique networks peak later, reflecting delayed inter-clique transmission.

In summary, the networks agree on mean degree (by construction) and on broad monotonic trends with 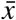, as well as on dynamics at 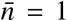 (where MC reduces to a random graph). They disagree sharply on clustering (high only in MC), on epidemic speed and intensity once 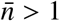 (MC slower, lower peaks, higher fade-out, delayed take off), and on the modulating effect of clique size (suppressive and delaying only in MC). These differences persist across stochastic realisations and demonstrate that explicit recurrent group structure with deterministic intra-group saturation and constrained inter-group pathways produces qualitatively and quantitatively distinct epidemic behaviour not reproduced by degree-matched ER, CM, or SBM networks.

## Discussion

The results (Figures 2–8 in the main text and the Supplementary figures) demonstrate that Multi-Clique networks produce systematically distinct epidemic dynamics compared to degree-matched Erdős–Rényi, configuration-model, and stochastic block model networks.

This structural slowdown arises from rapid saturation within the seed clique and constrained inter-clique escape via sparse external edges 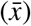, leading to delayed global takeoff and frequent early extinction. In contrast, ER and CM networks allow rapid, unimpeded spread across the entire graph once the degree distribution is fixed. This enables infection to explore the network more efficiently, leading to faster epidemic growth and higher peak prevalence than observed in MC networks. All models converge at 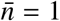 (where MC reduces to a random graph) and at high 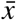, confirming that differences stem from the propensity to meet with everyone within cliques rather than degree or modularity alone. These findings align with prior work showing that strong local clustering and contact repetition limit transmission efficiency and raise effective epidemic thresholds compared to unclustered or random-mixing approximations [13, 22, 15].

We introduced the MC network as a *control-oriented* representation of recurrent physical contacts, explicitly separating deterministic within-group saturation from tunable between-group mixing. The external connectivity parameter 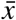 governs inter-clique transmission intensity and acts as a tunable lever for epidemic outcomes, even with fixed mean degree. Our simulations show high sensitivity to 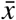: modest reductions suppress global spread and elevate fade-out probability (Figure 5), often causing outbreaks to self-extinguish within the initial clique without macroscopic propagation. The findings support known results indicating that classical epidemic models overestimate epidemic outcomes [12]. This places the MC network framework in a position to predict epidemic outcomes more realistically.

From a public health perspective, these results are important. Slower growth and higher fade-out captured by the MC network can enlarge the window for detection, surveillance, contact tracing, and early intervention before widespread transmission. Interventions targeting 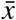 map directly to realistic measures—reducing inter-clique interactions while preserving within-team contacts, for example, limiting cross-class mixing in schools, or restricting inter-household visitation while maintaining household routines, and preserving essential social/economic activity while selectively weakening key transmission pathways. This contrasts with uniform contact-reduction strategies, which are often costly, less sustainable, and less effective in group-structured settings [2, 11].

The epidemiological implications of this structural bottleneck are substantial: classical models lacking explicit recurrent multi-clique group structure systematically overestimate speed, peak burden, and final size in populations with households, classrooms, or workplaces [13, 15]. MC networks capture the saturation and delayed inter-group spread observed in empirical contact data [17], suggesting improved realism for forecasting in such settings.

The MC framework is particularly data-friendly: parameters 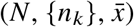 are estimable from routinely available sources—census/school records for group sizes/distributions, contact surveys/diaries for recurrent external contacts—without full network reconstruction or latent inference [17]. Interventions altering inter-group mixing map transparently onto 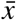, enabling quantitative scenario analysis aligned with policy granularity.

It is important to emphasise that observed differences do not arise from temporal variability—all models use static networks representing persistent opportunities. The distinguishing feature is the *organisation* of recurrent contacts into saturated, stable groups with limited inter-group connectivity, which characterises real-world physical interactions (households, classrooms, workplaces) but is homogenised in ER/CM or probabilistically relaxed in SBM.

By bridging abstract network theory and actionable strategy, MC offers a parsimonious yet expressive platform for evaluating structure-targeted interventions. Adoption of MC-like models could enhance forecast accuracy in group-structured populations, strengthen evidence for targeted controls, and support more precise, less disruptive public health responses.

### Scope and limitations

The Multi-Clique (MC) model targets epidemic spread in settings dominated by recurrent contacts within stable, persistent groups (e.g., households, classrooms, workplaces), where near-complete intra-group exposure and constrained inter-group mixing are key features. It may not be suitable for transient, one-off, or highly dynamic contact patterns, where temporal networks [10] or activity-driven models [21] are more appropriate.

This study is based entirely on stochastic, discrete-time SIR simulations on static MC networks. Except for the derivation of the mean degree, no analytical properties have been explored—including the epidemic threshold, basic reproduction number *R*_0_, critical phenomena, asymptotic outbreak size, or mean-field approximations—are presented here. These important theoretical aspects of the MC model are explicitly deferred to future analytical work, which could provide exact conditions for percolation, phase transitions, or comparisons to household-hybrid thresholds.

The model further assumes static, undirected edges without temporal ordering, edge turnover, or adaptive behaviour during outbreaks. While this aligns with persistent contact opportunities in many close-contact diseases, extensions to dynamic or adaptive networks could enhance realism in future studies.

Despite these limitations, the MC framework offers a data-friendly, interpretable abstraction that captures group-level saturation and bottlenecks more directly than standard random-graph or probabilistic block models. We hope that epidemic modellers would look into this direction, as this framework offers a physical contact network analogy relevant to and motivated by respiratory virus dissemination, thereby making it particularly useful for simulation-based policy exploration in structured populations.

## Data Availability

The R code for the MC network generation and the model simulation can be accessed through the link https://doi.org/10.5281/zenodo.21879408

## Ethics Declaration

This study focuses on the theoretical development of a mathematical model for the spread of a hypothetical infectious disease. No data was collected and analysed.

## Author contributions

Conceptualisation: M. L. S. Methodology: M. L. S. Supervision: A.S. & K. S. R. Software: K. S. R. & M. L. S. Visualisation: K S. R & M. L. S. Writing – original draft: M. L. S.

Writing – review & editing: A. S., K. S. R. & M. L. S.

