## Supplementary Information for "Recurrent Physical Contact Network Relevant for Epidemic Spreading"

Michael Lazarus Smah[1,2,*], Anna C. Seale[1,3], Kat S. Rock[2,4]

[1]Institute for Global Pandemic Planning, Warwick Medical School, Coventry, CV4 7AL, United Kingdom

[2]System Biology and Infectious Disease Epidemiology Research (SBIDER), University of Warwick, Coventry, CV4 7AL, United Kingdom

[3]London School of Hygiene & Tropical Medicine, London, United Kingdom

[4]Warwick Mathematics Institute, Coventry, CV4 7AL, United Kingdom

[*]Corresponding author: Michael Lazarus Smah


**Algorithm 1** Multi-Clique Network Construction

**Require:** Number of nodes $N$, mean clique size $\bar{n} > 1$, mean external degree $\bar{x} \geq 0$, core-node proportion $p_c \in (0, 1]$
**Ensure:** Undirected graph $G = (V, E)$ and adjacency matrix $A \in \{0, 1\}^{N \times N}$

Initialise node set $V = \{1, \ldots, N\}$ and edge set $E = \emptyset$
**Clique size generation**
Sequentially generate clique sizes $S_k = \max\{1, \tilde{S}_k\}$, where $\tilde{S}_k \sim \text{Poisson}(\bar{n})$, until all $N$ nodes have been allocated (i.e., $\sum_{k=1}^{K} S_k = N$).
**Node assignment**
Randomly permute the nodes and assign them to disjoint cliques $C_1, \ldots, C_K$ with sizes $S_1, \ldots, S_K$.
Store clique labels $\ell_u = k$ for each node $u \in C_k$.
**Intra-clique edges**
**for** each clique $C_k$ **do**
    **for** all distinct nodes $u, v \in C_k$ **do**
        Add edge $(u, v)$ to $E$.
    **end for**
**end for**
**External-degree targets**
Select $N_c = \lceil p_c N \rceil$ core nodes uniformly at random.
Assign initial external-degree targets $x_u$ such that $\frac{1}{N} \sum_{u \in V} x_u = \bar{x}$ and $\sum_{u \in V} x_u$ is even.
**Inter-clique pairing**
Create a stub list containing node $u$ repeated $x_u$ times.
Randomly shuffle the stub list.
**while** at least two stubs remain **do**
    Select a stub corresponding to node $u$.
    Identify candidate stubs corresponding to nodes $v$ satisfying $\ell_u \neq \ell_v$.
    **if** at least one candidate exists **then**
        Select a candidate uniformly at random.
        Add edge $(u, v)$ if it is not already present.
        Remove the paired stubs.
    **else**
        Remove the stub corresponding to $u$.
    **end if**
**end while**
**Isolation repair**
**for** each node $u$ with degree 0 **do**
    Select a node $v$ from a different clique.
    Add edge $(u, v)$.
**end for**
**Clique-connectivity repair**
**for** each clique $C_k$ **do**
    **if** $C_k$ has no external edge and $K > 1$ **then**
        Select a node $u \in C_k$.
        Select a node $v$ from another clique.
        Add edge $(u, v)$.
    **end if**
**end for**
**Adjacency matrix construction**
Construct the symmetric adjacency matrix $A$ from $E$.
**return** $G = (V, E)$ and $A$.

**Algorithm 1** is a pseudocode, which provides a procedure for generating realisations of the Multi-Clique (MC) network model.

### Results

Figures 1–6 in this supplementary information show that degree distributions are well-matched across models. For every $(\bar{n}, \bar{x})$ pair the boxplots of realised mean degree show overlapping distributions and similar medians/variability for ER, CM, MC, and SBM, with differences falling below one, except that the mean degree of CM is higher for $\bar{n} = 1, 2$. Minor stochastic differences exist (especially at low $\bar{n}$), but the matching procedure succeeds. At $\bar{n} = 1$ the MC model collapses to a degree-controlled random graph and all four overlap almost completely, however, as $\bar{n}$ is increased, intra-clique contributions $(\bar{n} - 1)$ increase $\langle k \rangle$ predictably for all realisations. Hence, all models are matched, except for the structural variability, upon which epidemic outcomes would rely.

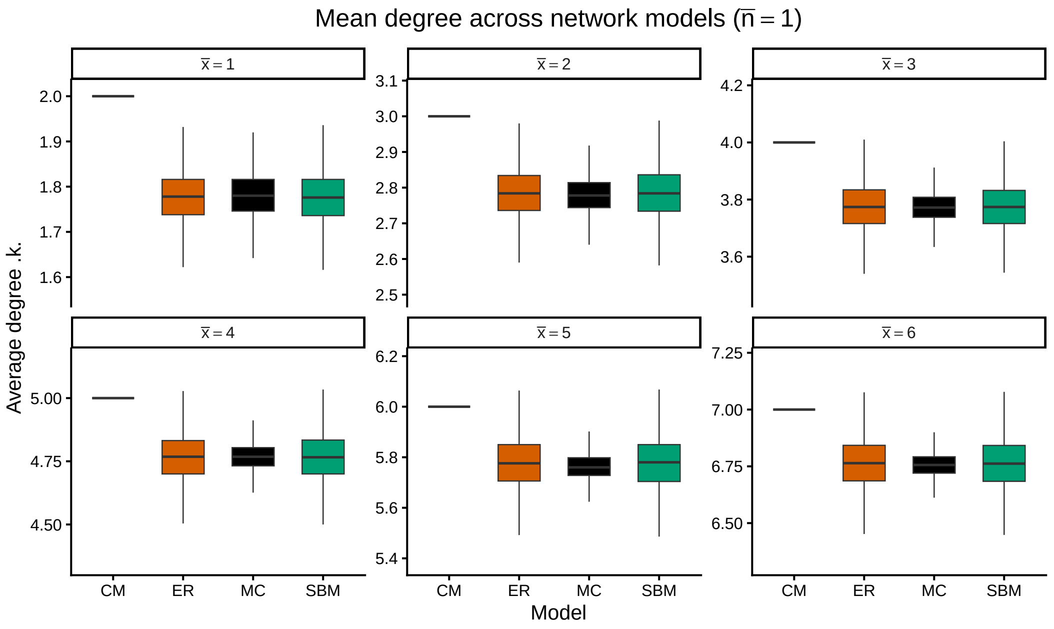


Figure 1: Mean degree distributions for Erdős–Rényi (ER), stochastic block model (SBM), configuration model (CM), and multi-clique (MC) networks at average clique size $\bar{n} = 1$. Each panel fixes external connectivity $\bar{x}$, numbered 1–6 at the top of each panel.

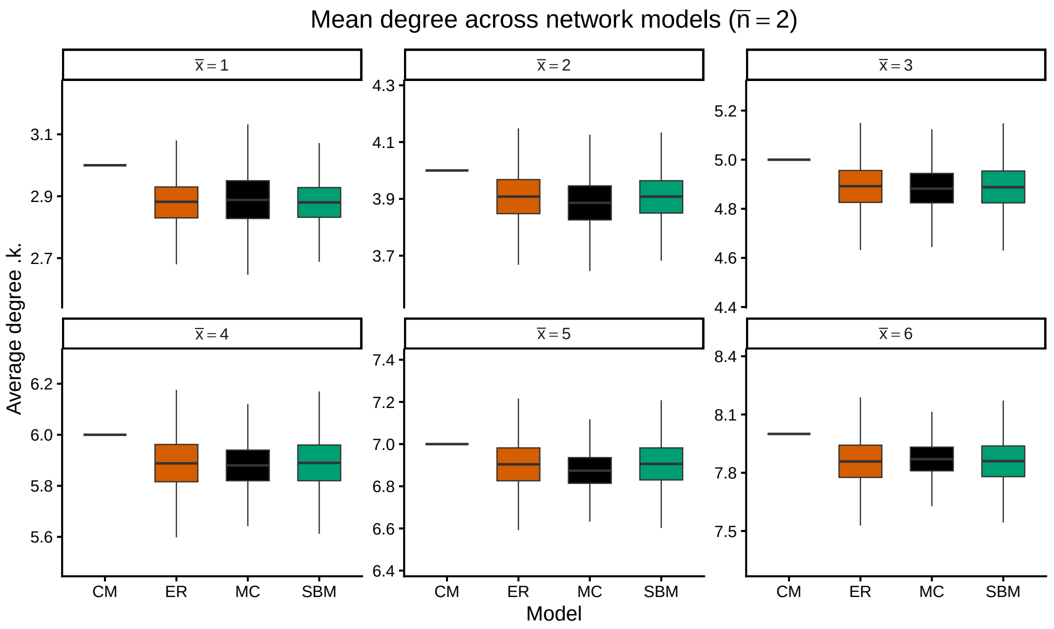


Figure 2: Mean degree distributions for Erdős–Rényi (ER), stochastic block model (SBM), configuration model (CM), and multi-clique (MC) networks at average clique size $\bar{n} = 2$. Each panel fixes external connectivity $\bar{x}$, numbered 1–6 at the top of each panel.

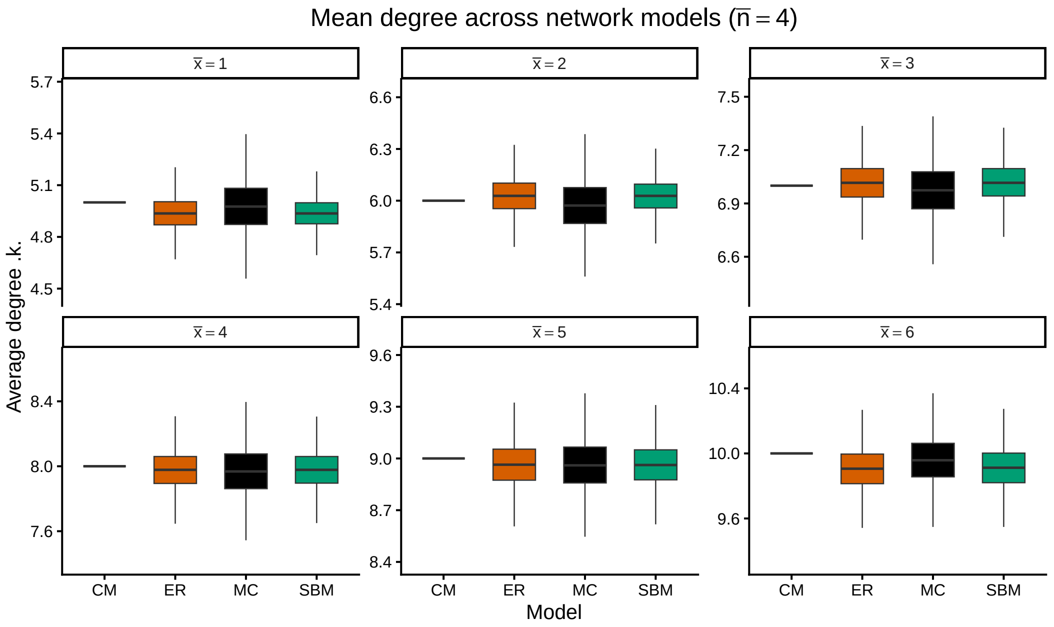


Figure 3: Mean degree distributions for Erdős–Rényi (ER), stochastic block model (SBM), configuration model (CM), and multi-clique (MC) networks at average clique size $\bar{n} = 4$. Each panel fixes external connectivity $\bar{x}$, numbered 1–6 at the top of each panel.

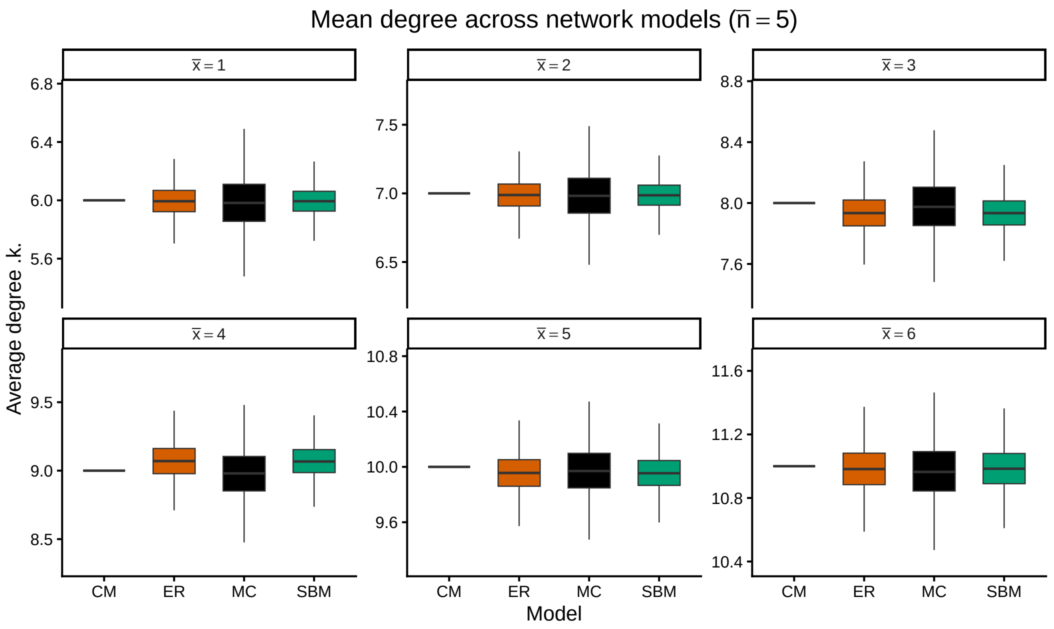


Figure 4: Mean degree distributions for Erdős–Rényi (ER), stochastic block model (SBM), configuration model (CM), and multi-clique (MC) networks at average clique size $\bar{n} = 5$. Each panel fixes external connectivity $\bar{x}$, numbered 1–6 at the top of each panel.

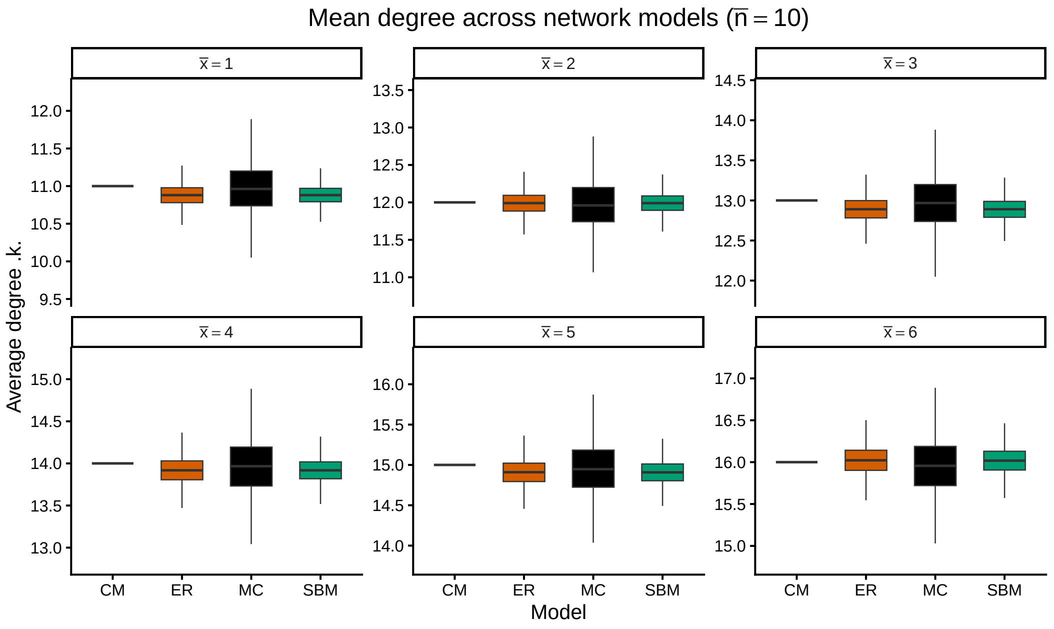


Figure 5: Mean degree distributions for Erdős–Rényi (ER), stochastic block model (SBM), configuration model (CM), and multi-clique (MC) networks at average clique size $\bar{n} = 10$. Each panel fixes external connectivity $\bar{x}$, numbered 1–6 at the top of each panel.

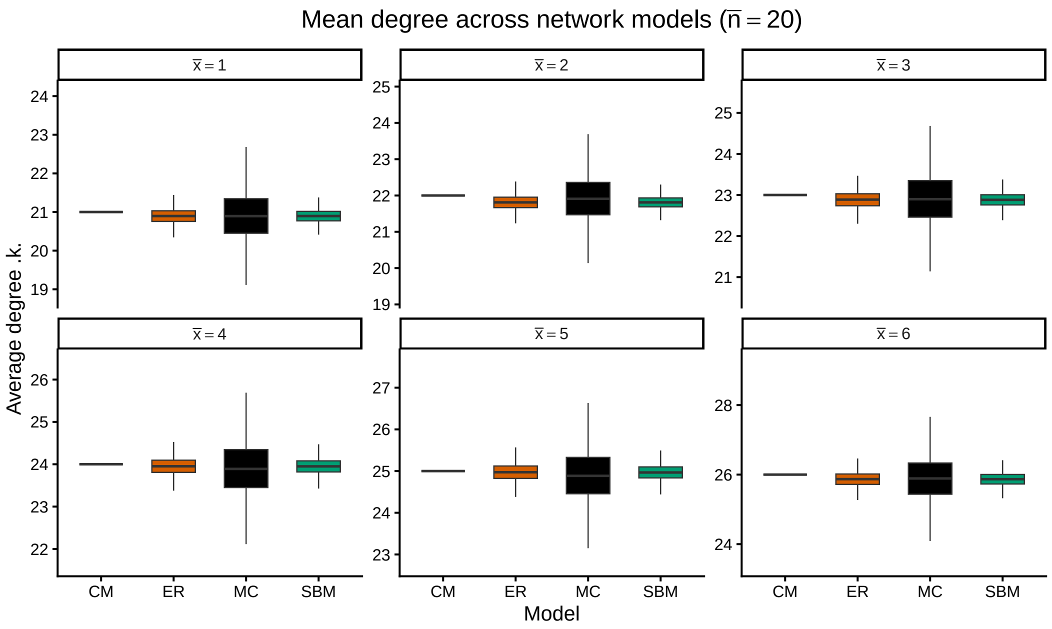


Figure 6: Mean degree distributions for Erdős–Rényi (ER), stochastic block model (SBM), configuration model (CM), and multi-clique (MC) networks at average clique size $\bar{n} = 20$. Each panel fixes external connectivity $\bar{x}$, numbered 1–6 at the top of each panel.

Figures 7–12 showing local clustering reveals the core structural distinction. MC networks exhibit substantially higher average local clustering coefficients than the comparator-models for all $\bar{n} > 1$, with the gap widening dramatically as $\bar{n}$ increases. For $\bar{n} = 1$ clustering is low ($\approx 0$) and comparable across all four models. For $\bar{n} = 2$ clustering in MC is already markedly elevated; by $\bar{n} = 5$ it reaches 0.4–0.6 depending on $\bar{x}$, and for $\bar{n} = 10$–20 it approaches 1. In contrast, ER, CM, and SBM clustering remains near zero (typically $< 0.05$) across the entire range of $\bar{n}$ and $\bar{x}$. Within MC, clustering decreases with increasing $\bar{x}$ once $\bar{n}$ is fixed.

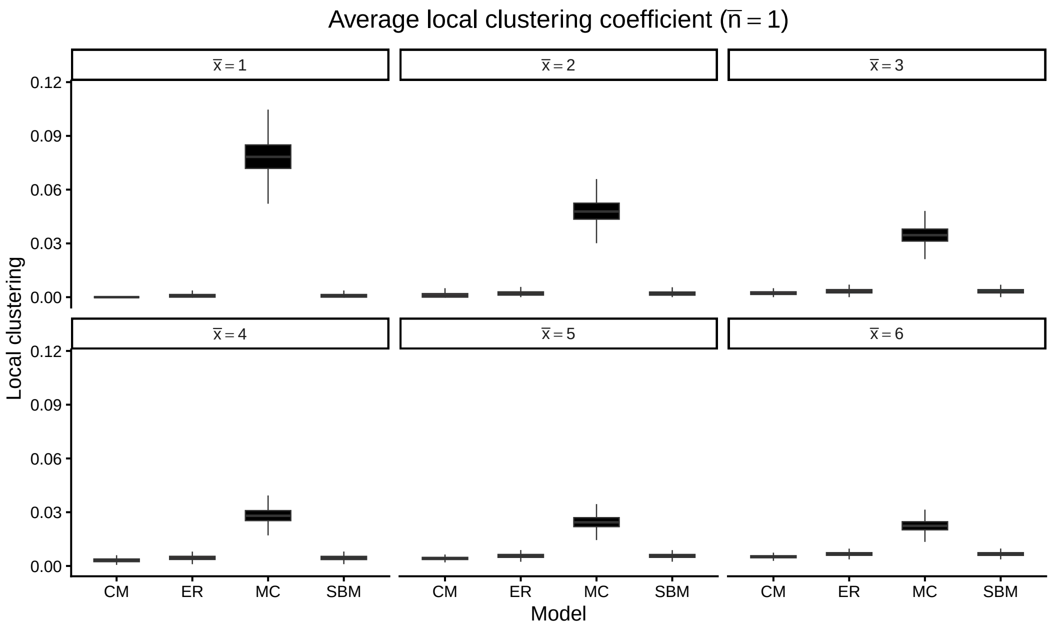


Figure 7: Average local clustering coefficient across network models for $\bar{n} = 1$ and varying external connectivity $\bar{x}$. Multi-clique networks exhibit substantially higher clustering than degree-matched comparing models.

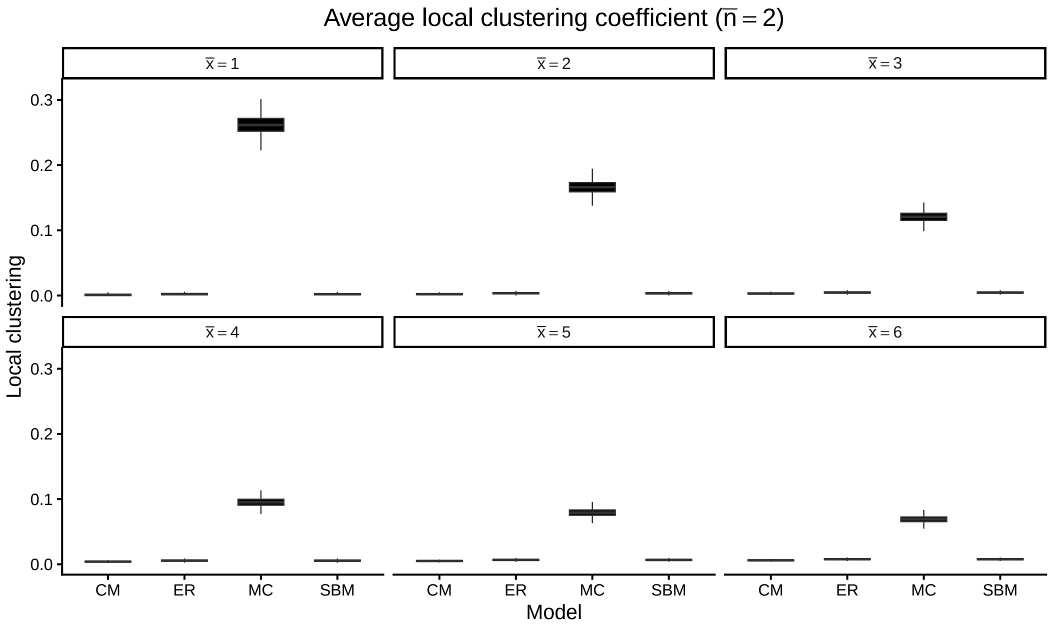


Figure 8: Average local clustering coefficient across network models for $\bar{n} = 2$ and varying external connectivity $\bar{x}$. Multi-clique networks exhibit substantially higher clustering than degree-matched comparing models.

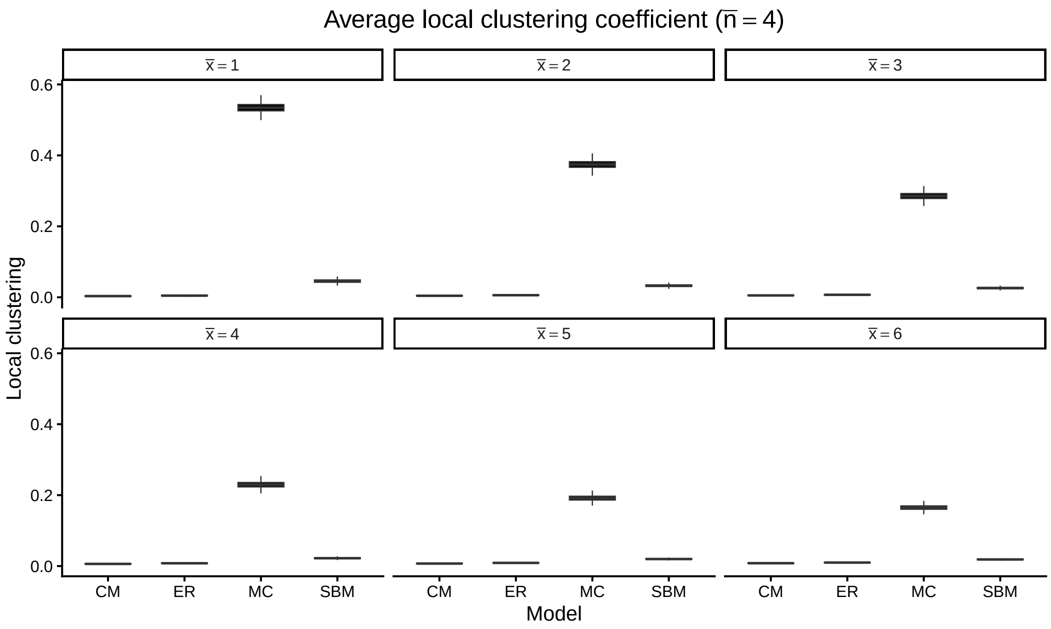


Figure 9: Average local clustering coefficient across network models for $\bar{n} = 4$ and varying external connectivity $\bar{x}$. Multi-clique networks exhibit substantially higher clustering than degree-matched comparing models.

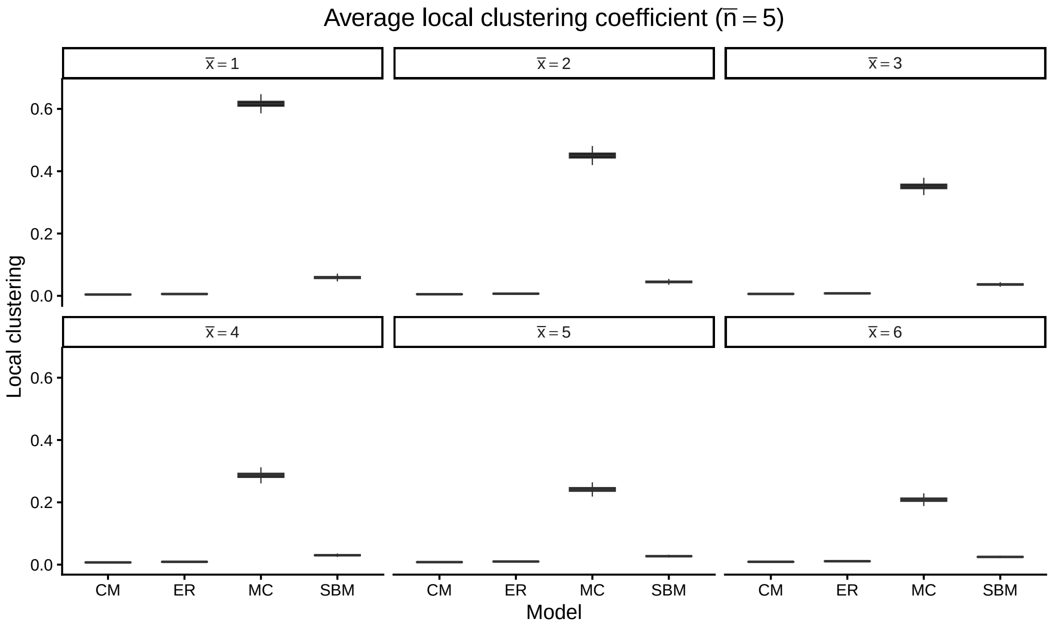


Figure 10: Average local clustering coefficient across network models for $\bar{n} = 5$ and varying external connectivity $\bar{x}$. Multi-clique networks exhibit substantially higher clustering than degree-matched comparing models.

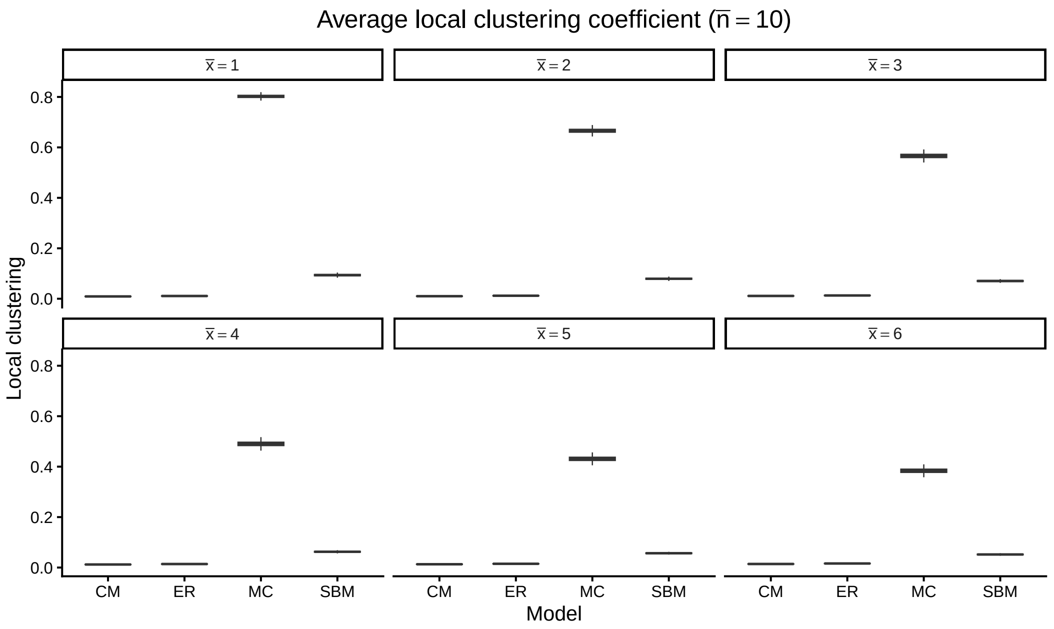


Figure 11: Average local clustering coefficient across network models for $\bar{n} = 10$ and varying external connectivity $\bar{x}$. Multi-clique networks exhibit substantially higher clustering than degree-matched comparing models.

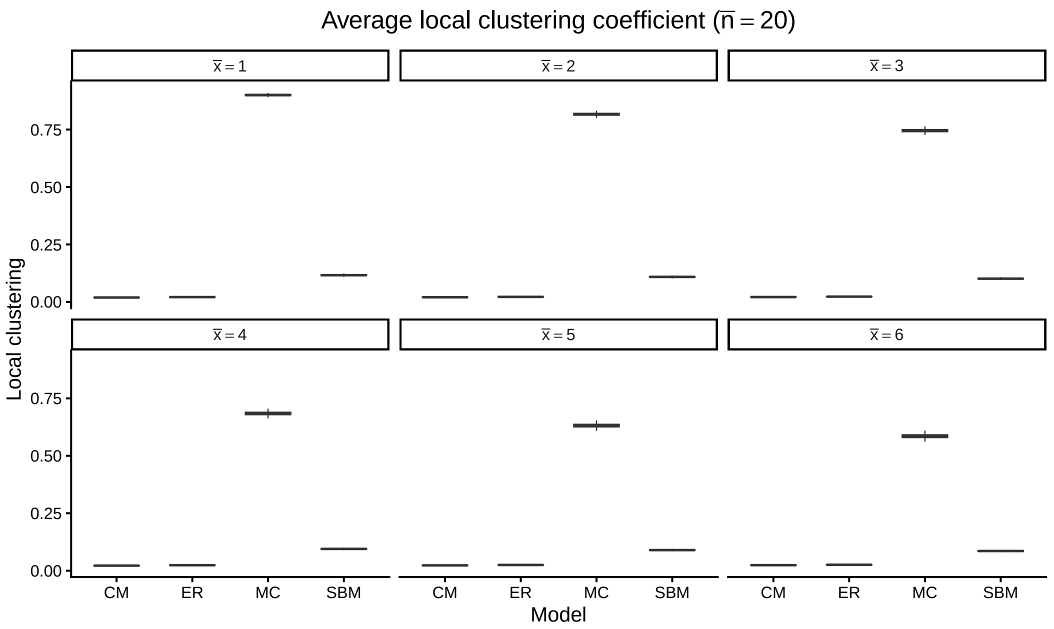


Figure 12: Average local clustering coefficient across network models for $\bar{n} = 20$ and varying external connectivity $\bar{x}$. Multi-clique networks exhibit substantially higher clustering than degree-matched comparing models.

Figures 13–18, 19–24 show that epidemic trajectories differ systematically once clique structure is present. At $\bar{n} = 1$ all four models produce nearly indistinguishable mean infection curves for every $\bar{x}$: rapid rise, similar peak height and timing, and comparable decay. As $\bar{n}$ increases the MC trajectories diverge. For $\bar{n} \geq 4$–5 (especially at low-to-moderate $\bar{x}$) MC outbreaks show delayed take off, lower peak prevalence, and slower overall growth relative to ER, CM, and SBM. The delay is most pronounced at small $\bar{x}$, where infection often saturates the initial clique before inter-clique transmission occurs; higher $\bar{x}$ allows earlier escape but MC still lags the benchmarks. ER and CM curves remain fast and similar to each other; SBM is intermediate but faster than MC because probabilistic within-block mixing avoids full local saturation. At high $\bar{x}$ (5–6) and large $\bar{n}$ the qualitative shapes converge somewhat, but MC peaks remain delayed. When plotted per network type (Figs. 19–24), MC curves at low $\bar{x}$ are visibly flattened and right-shifted compared with the overlapping families of curves for the other three models.

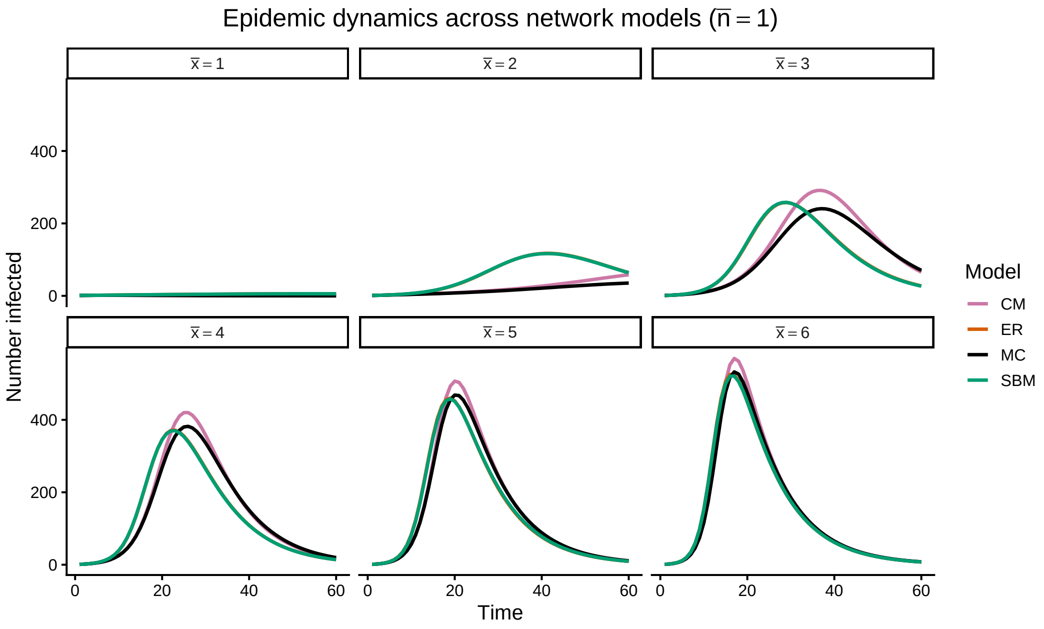


Figure 13: Network structure shapes epidemic trajectories. Mean SIR infection dynamics for $\bar{n} = 1$ across network models. Each panel fixes external connectivity $\bar{x}$. Curves show averages over stochastic simulations.

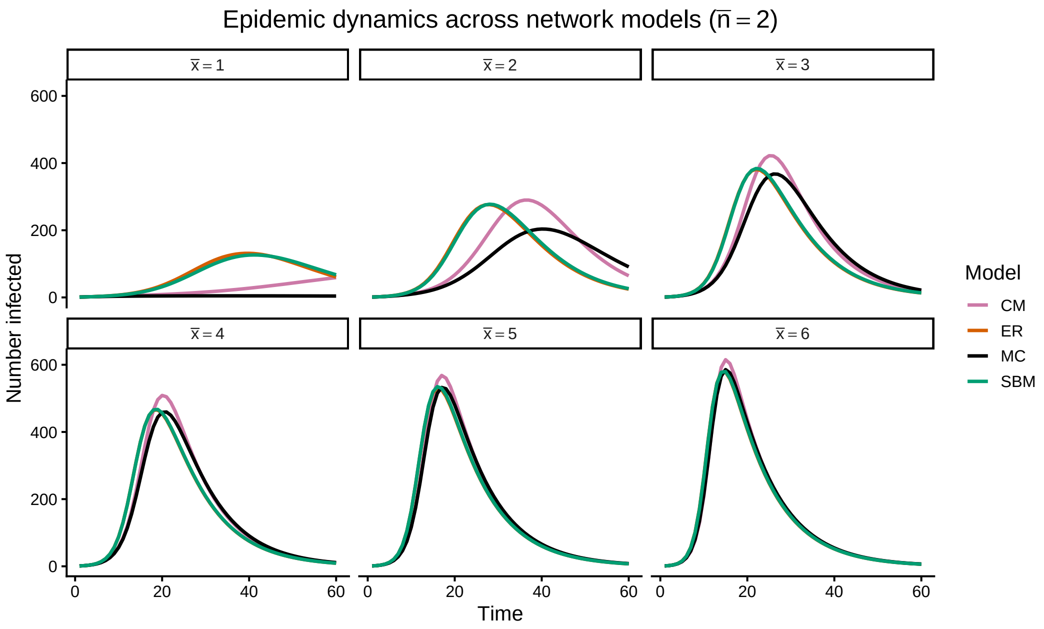


Figure 14: Mean SIR infection dynamics for $\bar{n} = 2$ across network models. Each panel fixes external connectivity $\bar{x}$. Curves show averages over stochastic simulations.

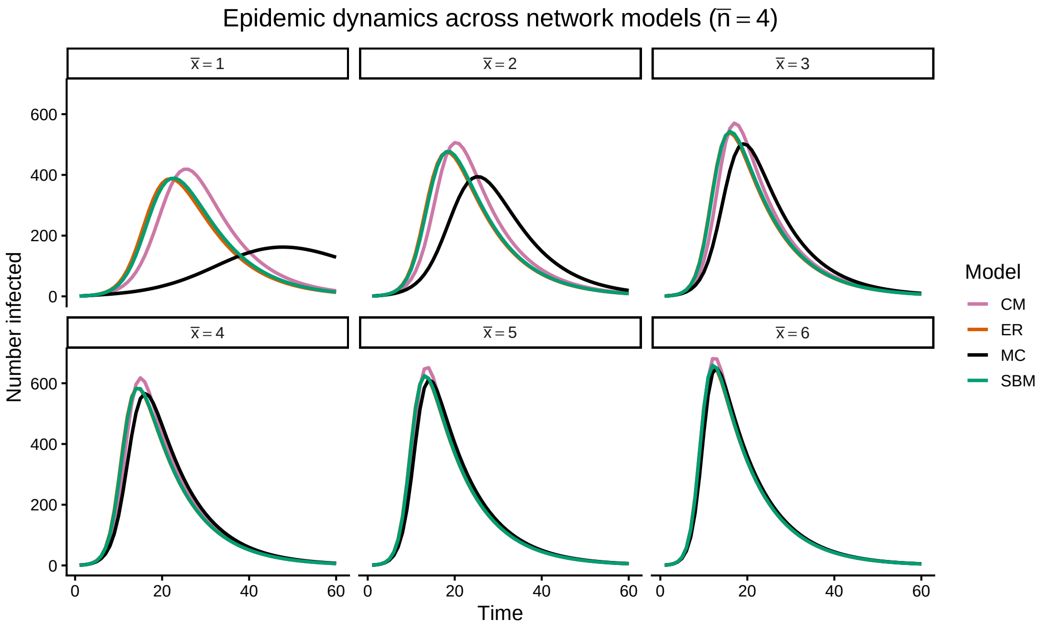


Figure 15: Mean SIR infection dynamics for $\bar{n} = 4$ across network models. Each panel fixes external connectivity $\bar{x}$. Curves show averages over stochastic simulations.

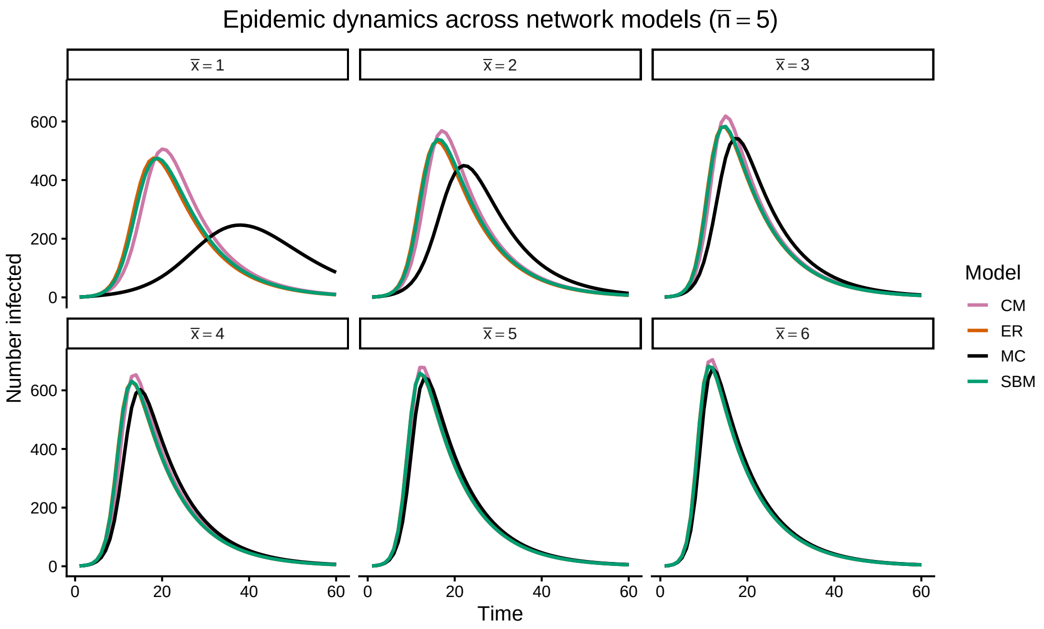


Figure 16: Mean SIR infection dynamics for $\bar{n} = 5$ across network models. Each panel fixes external connectivity $\bar{x}$. Curves show averages over stochastic simulations.

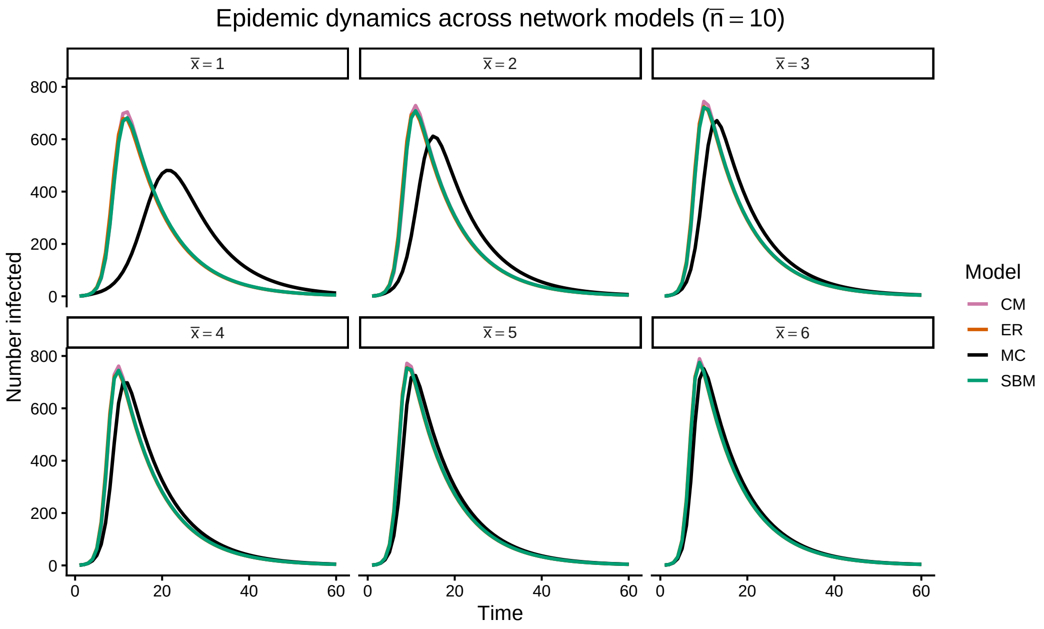


Figure 17: Mean SIR infection dynamics for $\bar{n} = 10$ across network models. Each panel fixes external connectivity $\bar{x}$. Curves show averages over stochastic simulations.

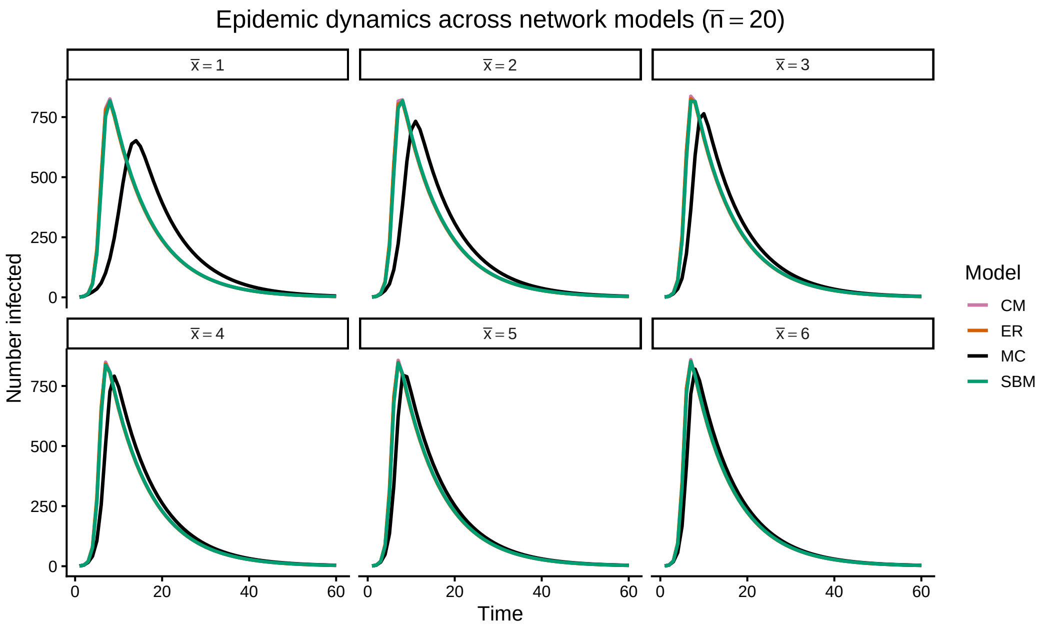


Figure 18: Mean SIR infection dynamics for $\bar{n} = 20$ across network models. Each panel fixes external connectivity $\bar{x}$. Curves show averages over stochastic simulations.

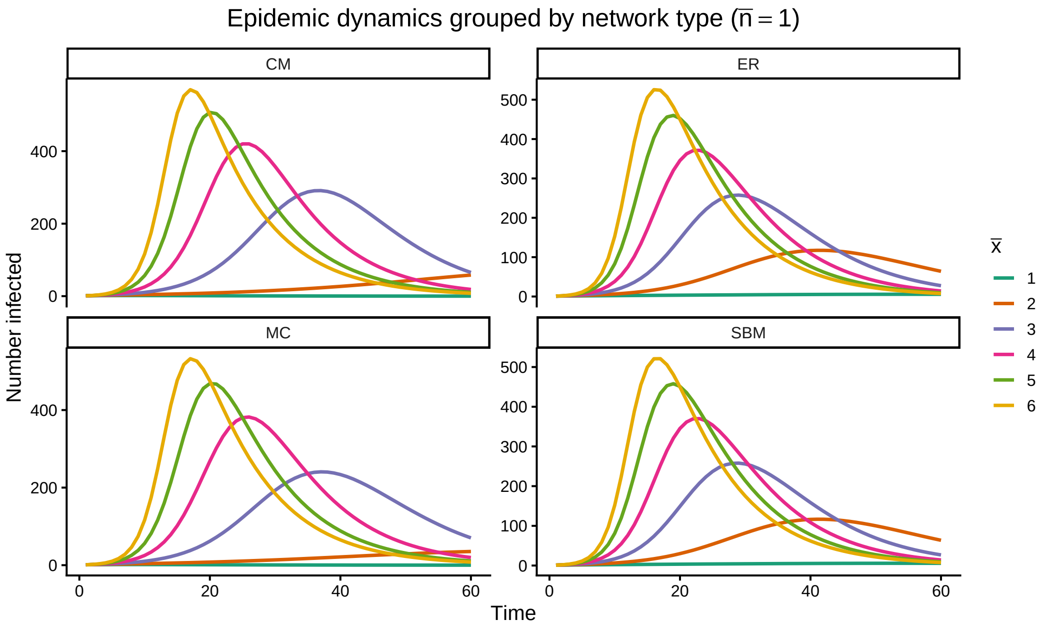


Figure 19: Qualitative epidemic dynamics by network architecture. Mean infection trajectories for $\bar{n} = 1$, shown separately for each network model. Colours indicate external connectivity $\bar{x}$. Multi-clique networks display delayed and attenuated outbreaks at low $\bar{x}$, reflecting strong local clustering.

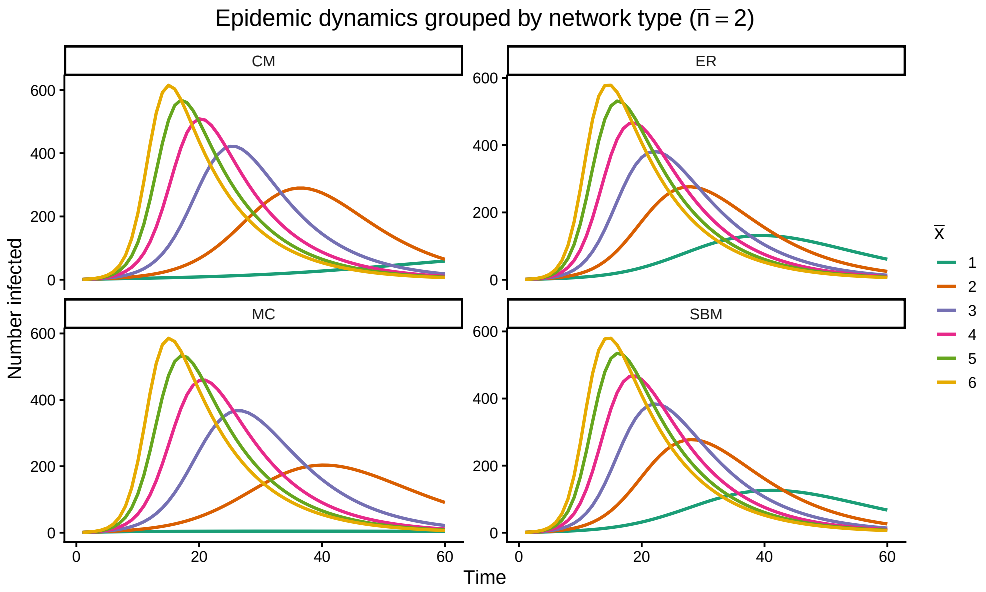


Figure 20: Qualitative epidemic dynamics by network architecture. Mean infection trajectories for $\bar{n} = 2$, shown separately for each network model. Colours indicate external connectivity $\bar{x}$. Multi-clique networks display delayed and attenuated outbreaks at low $\bar{x}$, reflecting strong local clustering.

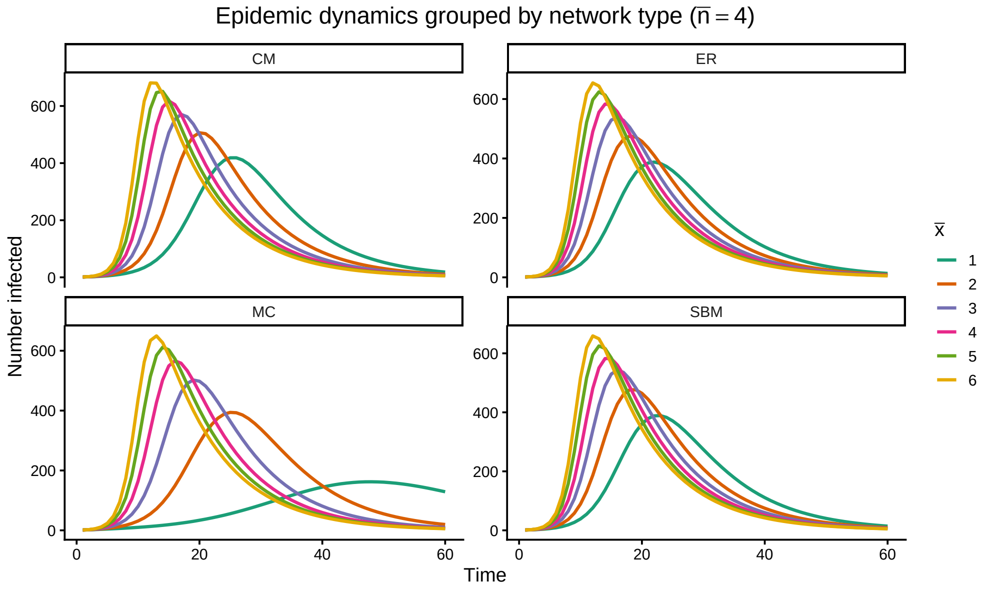


Figure 21: Qualitative epidemic dynamics by network architecture. Mean infection trajectories for $\bar{n} = 4$, shown separately for each network model. Colours indicate external connectivity $\bar{x}$. Multi-clique networks display delayed and attenuated outbreaks at low $\bar{x}$, reflecting strong local clustering.

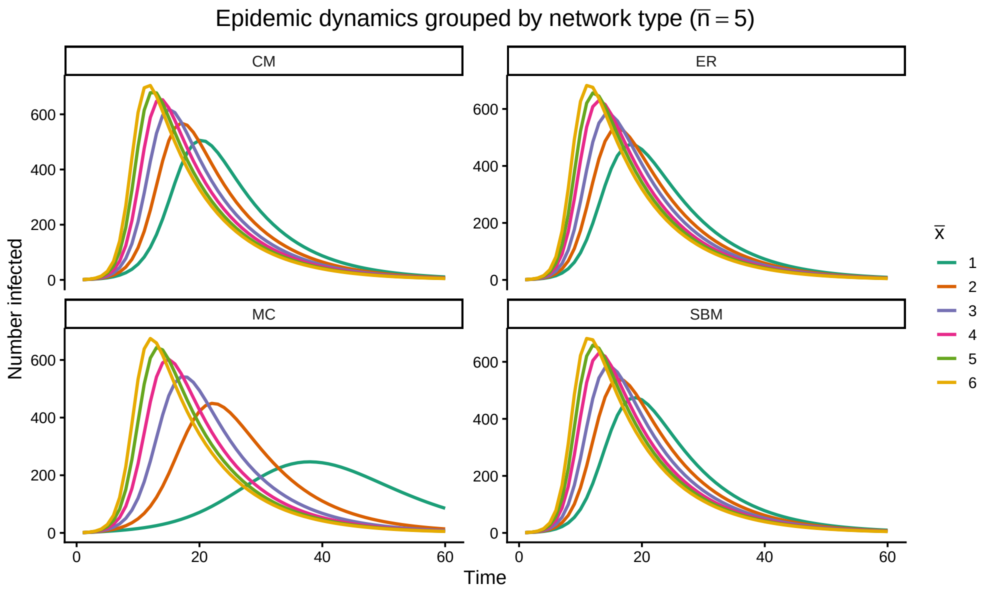


Figure 22: Qualitative epidemic dynamics by network architecture. Mean infection trajectories for $\bar{n} = 5$, shown separately for each network model. Colours indicate external connectivity $\bar{x}$. Multi-clique networks display delayed and attenuated outbreaks at low $\bar{x}$, reflecting strong local clustering.

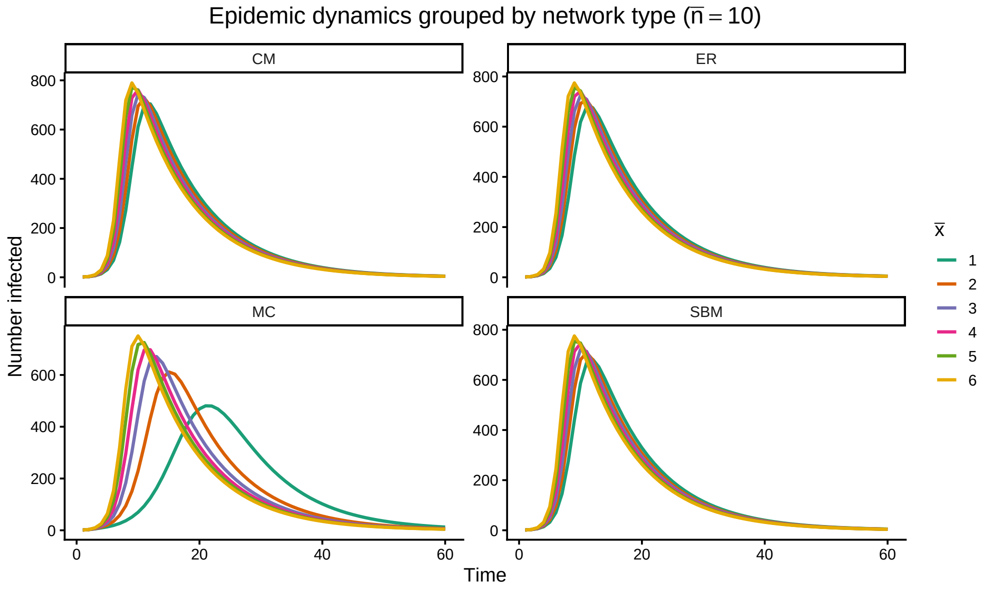


Figure 23: Qualitative epidemic dynamics by network architecture. Mean infection trajectories for $\bar{n} = 10$, shown separately for each network model. Colours indicate external connectivity $\bar{x}$. Multi-clique networks display delayed and attenuated outbreaks at low $\bar{x}$, reflecting strong local clustering.

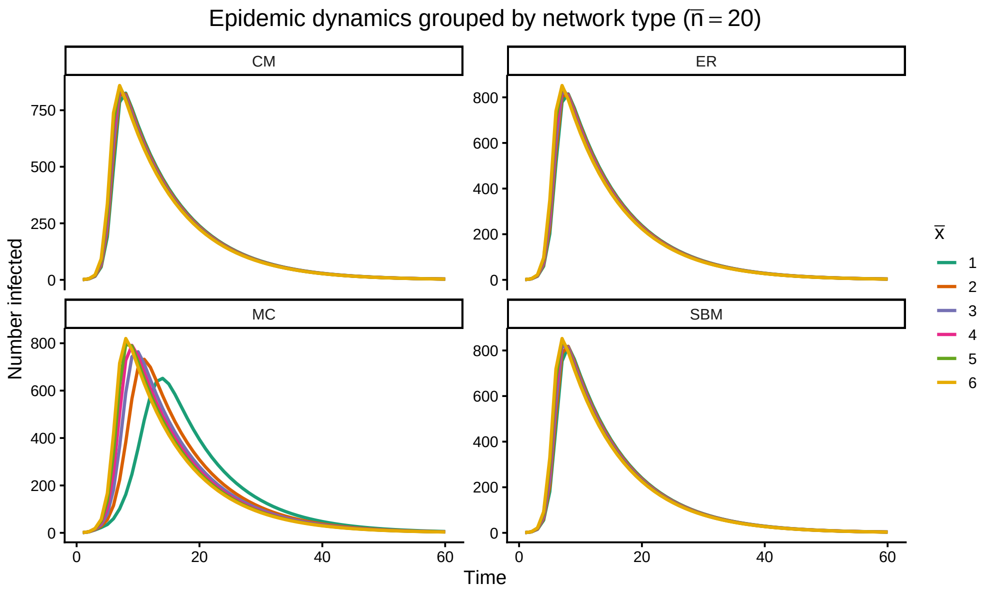


Figure 24: Qualitative epidemic dynamics by network architecture. Mean infection trajectories for $\bar{n} = 20$, shown separately for each network model. Colours indicate external connectivity $\bar{x}$. Multi-clique networks display delayed and attenuated outbreaks at low $\bar{x}$, reflecting strong local clustering.

Figures 25–30 showing summary epidemic outcomes confirm the structural effects of MC network. Final epidemic size increases monotonically with $\bar{x}$ in all models and approaches similar values at high $\bar{x}$, indicating agreement on ultimate reach once transmission percolates globally. However, MC consistently shows modestly lower final sizes at intermediate $\bar{x}$, especially for larger $\bar{n}$. Peak prevalence follows the same pattern: comparable at $\bar{n} = 1$ and high $\bar{x}$, but MC exhibits lower peaks (sometimes substantially) for $\bar{n} \geq 5$ and $\bar{x} \leq 4$. Fade-out probability is markedly higher on MC networks at low $\bar{x}$, reflecting frequent extinction inside the seed clique. The time to peak is longer on MC across almost all regimes, with the largest differences at small $\bar{x}$ and moderate-to-large $\bar{n}$; ER, CM, and SBM peak earlier. All models agree on the qualitative monotonicity (larger $\bar{x} \to$ bigger/faster outbreaks, lower fade-out, earlier peaks), but disagree quantitatively once $\bar{n} > 1$, with MC systematically slower, lower-peaking, and more prone to stochastic extinction.

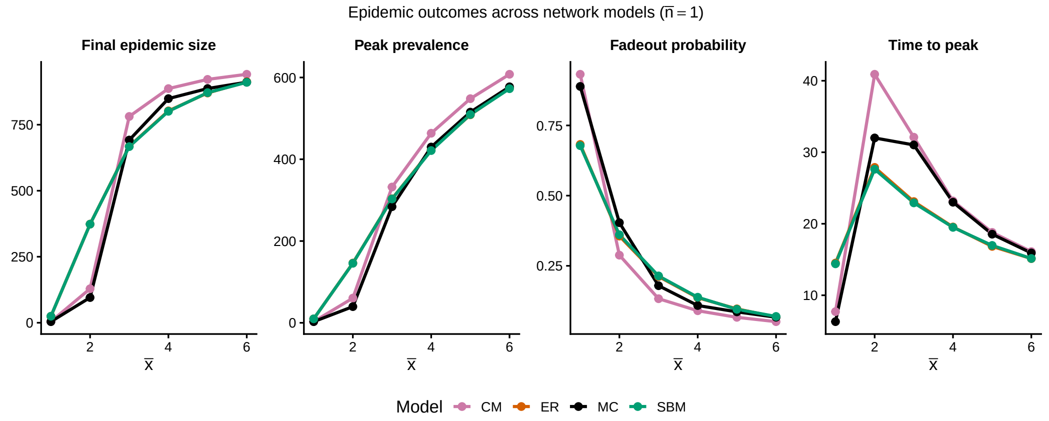


Figure 25: Summary of epidemic outcomes across network models. Final epidemic size, peak prevalence, fadeout probability, and time to peak as functions of external connectivity $\bar{x}$ for $\bar{n} = 1$. Values are averaged across 1,000 stochastic simulations.

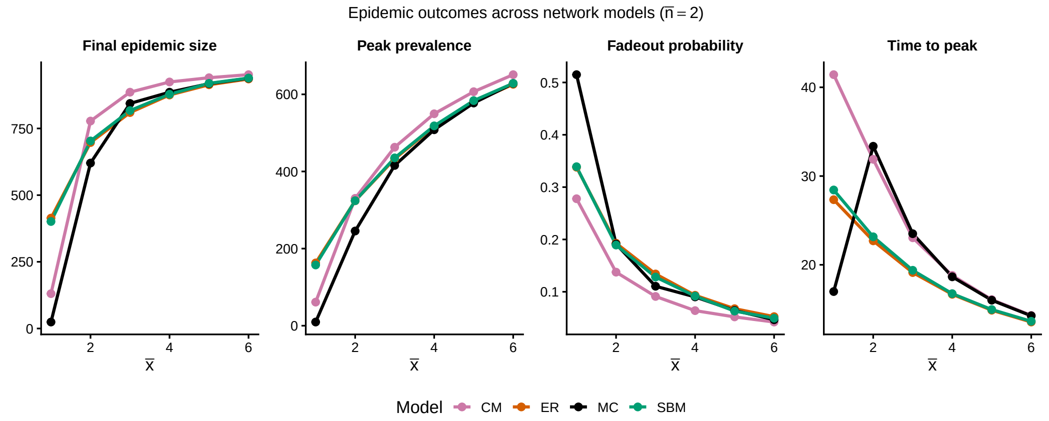


Figure 26: Summary of epidemic outcomes across network models. Final epidemic size, peak prevalence, fadeout probability, and time to peak as functions of external connectivity $\bar{x}$ for $\bar{n} = 2$. Values are averaged across 1,000 stochastic simulations.

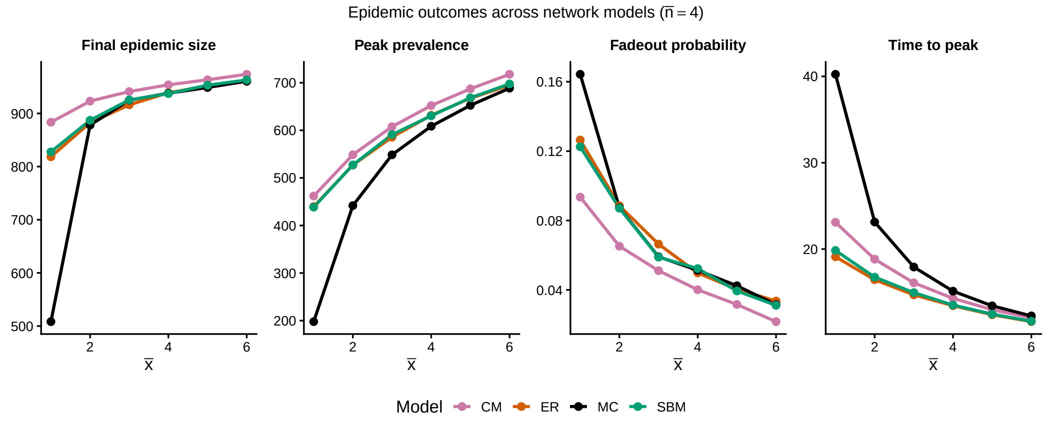


Figure 27: Summary of epidemic outcomes across network models. Final epidemic size, peak prevalence, fadeout probability, and time to peak as functions of external connectivity $\bar{x}$ for $\bar{n} = 4$. Values are averaged across 1,000 stochastic simulations.

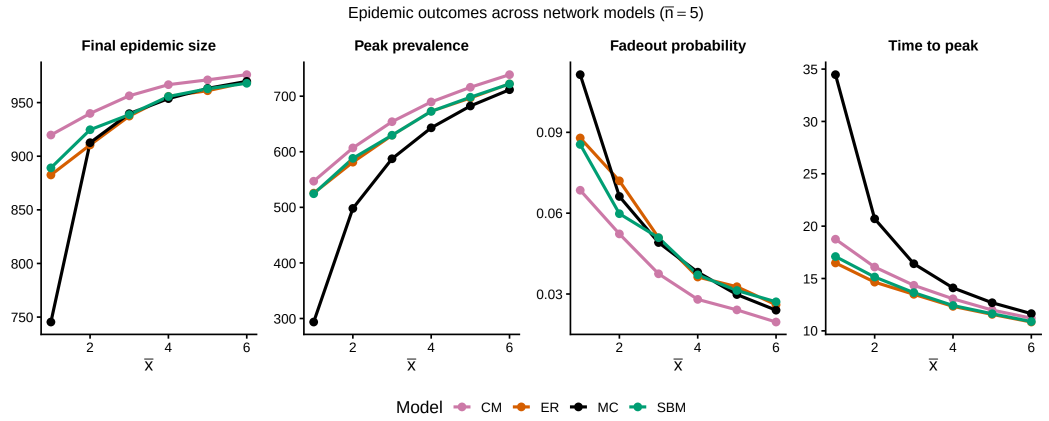


Figure 28: Summary of epidemic outcomes across network models. Final epidemic size, peak prevalence, fadeout probability, and time to peak as functions of external connectivity $\bar{x}$ for $\bar{n} = 5$. Values are averaged across 1,000 stochastic simulations.

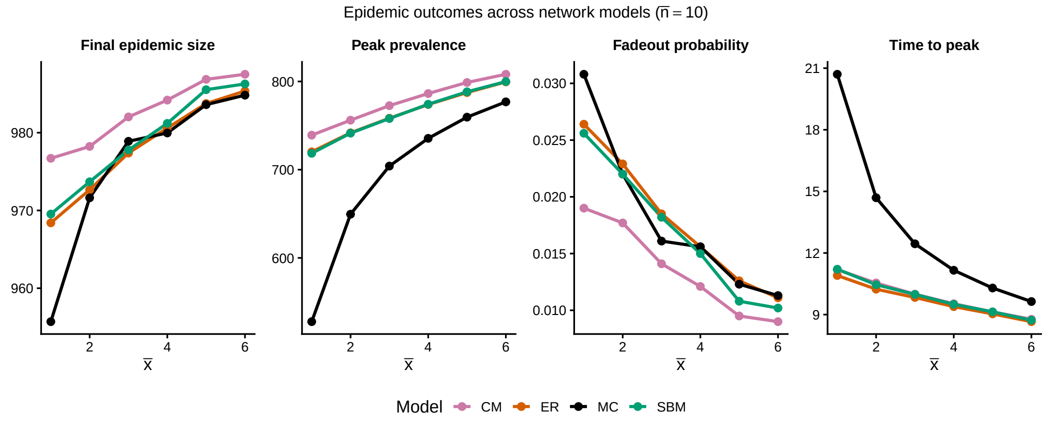


Figure 29: Summary of epidemic outcomes across network models. Final epidemic size, peak prevalence, fadeout probability, and time to peak as functions of external connectivity $\bar{x}$ for $\bar{n} = 10$. Values are averaged across 1,000 stochastic simulations.

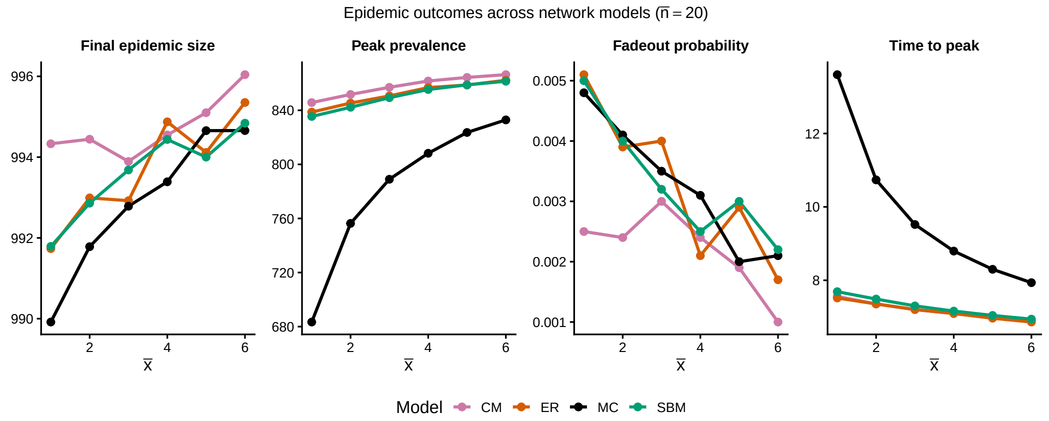


Figure 30: Summary of epidemic outcomes across network models. Final epidemic size, peak prevalence, fadeout probability, and time to peak as functions of external connectivity $\bar{x}$ for $\bar{n} = 20$. Values are averaged across 1,000 stochastic simulations.

Figures 31–33 show suppressed and delayed epidemics in MC networks relative to the comparator models, and the differences become more pronounced with increasing the mean clique size $\bar{n}$. For fixed $\bar{x}$, final epidemic size declines mildly with $\bar{n}$ on MC while remaining high and stable on ER/CM/SBM; the relative gap widens at lower $\bar{x}$. Peak prevalence decreases more noticeably with $\bar{n}$ on MC than on the benchmarks, which maintain higher peaks. Time to peak shortens overall with $\bar{n}$ (larger cliques accelerate local saturation), but MC still peaks later than the others for each fixed $\bar{n}$ and $\bar{x}$. These trends reinforce that deterministic full connectivity within larger groups creates strong local bottlenecks that the probabilistic mixing of SBM (and the lack of explicit groups in ER/CM) do not replicate.

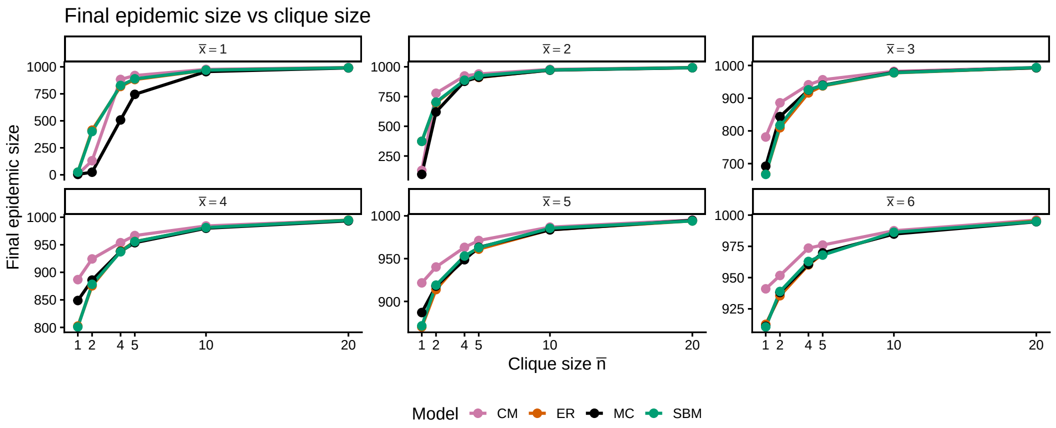


Figure 31: Final epidemic size as a function of average clique size $\bar{n}$. Each panel fixes external connectivity $\bar{x}$. Increasing $\bar{n}$ reduces outbreak size in multi-clique networks relative to degree-matched alternatives.

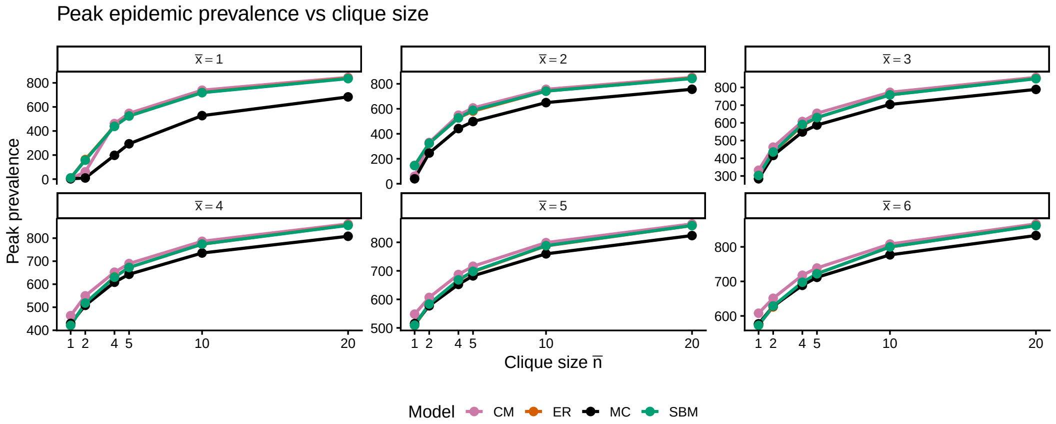


Figure 32: Peak epidemic prevalence as a function of average clique size $\bar{n}$ for fixed external connectivity $\bar{x}$. Multi-clique networks consistently exhibit lower peaks than comparing models.

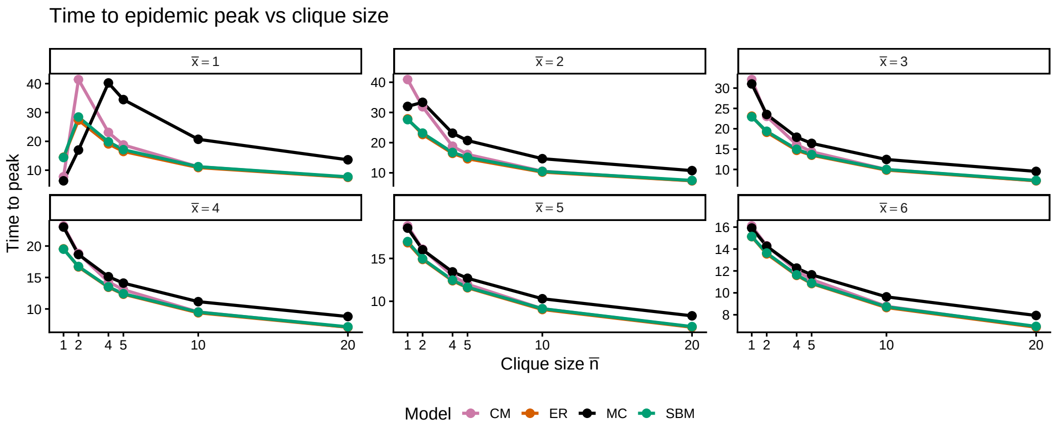


Figure 33: Time to epidemic peak as a function of average clique size $\bar{n}$. Each panel fixes external connectivity $\bar{x}$. Epidemics on multi-clique networks peak later, reflecting delayed inter-clique transmission.
